# Viral introductions and return to baseline sexual behaviors maintain low-level mpox incidence in Los Angeles County, USA, 2023-2024

**DOI:** 10.1101/2025.03.14.25323999

**Authors:** Miguel I. Paredes, Citina Liang, Sze-chuan Suen, Ian W. Holloway, Jacob M. Garrigues, Nicole M. Green, Trevor Bedford, Nicola F. Müller, Joseph Osmundson

## Abstract

In 2022, mpox clade IIb disseminated around the world, causing outbreaks in more than 117 countries. Despite the decay of the 2022 epidemic and the accumulation of immunity within queer sexual networks, mpox continues to persist at low incidence in North America without extinction, raising concerns of future outbreaks. We combined phylodynamic inference and microsimulation modeling to understand the heterogeneous dynamics governing local mpox persistence in Los Angeles County (LAC) from 2023-2024. Our Bayesian phylodynamic analysis revealed a time-varying pattern of viral importations into the county that seeded a skewed distribution of mpox outbreak clusters that display a “stuttering chains” dynamic. Our phylodynamics-informed microsimulation model demonstrated that the persistent number of mpox cases in LAC can be explained by a combination of waves of viral introductions, a median *Rt* significantly below one, and a return to near-baseline sexual behaviors that were altered during the 2022 epidemic. Finally, our counterfactual scenario modeling showed that public health interventions that either promote increased isolation of symptomatic, infectious individuals or enact behavior-modifying campaigns during the periods with the highest viral importation intensity are both actionable and effective at curbing mpox cases. Our work highlights the heterogeneous factors that maintain present-day mpox dynamics in a large, urban US county and describes how to leverage these results to design timely and community-centered public health interventions.

## Introduction

Mpox is a viral infection caused by the monkeypox virus (MPXV), an orthopoxvirus closely related to smallpox (1). In 2022, mpox spread globally, largely via queer sexual networks, causing tens of thousands of cases (2). Mpox clade IIb was introduced into humans around 2014 in Nigeria (3), and was the causative genetic clade of the 2022 outbreak. Mpox clade IIb continues to spread around the world, including in the United States (US) (4–6). Additionally, a current outbreak of clade I in Central Africa also raises concern of international spread (7,8).

In 2022, clade IIb mpox cases in the US reached over one hundred per day. Mpox infections in the US have since remained at low, but persistent, levels (9,10). While sporadic larger mpox outbreaks have occurred, they have neither grown to a large-scale epidemic nor been eradicated, as would be expected if the effective reproduction number (*Rt*) was above or below one, respectively (11). The mechanisms maintaining sporadic mpox incidence locally could include viral introductions via travel (12), small local clusters where *Rt* is larger than one (e.g. heavy-tailed infection dynamics) (13), a combination of these factors, or other undescribed mechanisms. If either travel or limited local clusters cause a majority of mpox transmission in a specific geographical location, targeted public health interventions that respond to the dynamics of the epidemic could potentially prevent a large proportion of mpox cases.

Disentangling the contribution of travel-related and local transmission on infectious disease dynamics is difficult from case counts alone. Alternatively, phylodynamics allows for tracking of viral movement across time and space via analysis of viral genomes (14). Prior work has employed phylodynamics to understand global, regional, and local mpox spread by leveraging global sequencing efforts to examine mpox transmission prior to widespread testing availability and to understand the interplay between viral introductions and local spread (6,15,16). Phylodynamics works in a retrospective fashion to model viral evolution and transmission.

Microsimulation models simulate individual-level disease trajectories by combining mechanistic descriptions of disease processes with empirical data and estimated parameters, both measured and estimated, to determine the dynamics underlying infectious disease transmission (17). By simulating counterfactual scenarios, microsimulations can elucidate the mechanistic factors that determine and curb spread. Prior microsimulation work on mpox dynamics has been used at a local level to both understand factors that promoted the decline of the 2022 epidemic and to test the effectiveness of public health interventions (18). Microsimulation models are, however, often limited by data availability and model assumptions.

To address these shortcomings, we combine phylodynamics and microsimulation modeling to understand mpox spread in Los Angeles County (LAC) in 2023 and 2024. We employ Bayesian phylodynamics to estimate mpox importation dynamics into LAC, and use our phylodynamic results to parameterize a microsimulation model of mpox with a force of viral importations. Through our combined approach, we estimate the role of various factors in promoting mpox persistence in 2023-2024, such as the return of baseline sexual behaviors, rates of isolation for those with diagnosed mpox, and the role of importation versus local mpox transmission. We then use our mpox microsimulation model to evaluate the public health impact of interventions that target the identified dynamics of local spread. The combination of these two computational methods serves to answer complex and essential questions regarding the largely understudied current state of the mpox epidemic and provides direct opportunities for public health action..

## Results

### Microsimulation shows local cases in LAC die out in 2023-2024 without viral importations

After mpox was initially detected in May 2022, the number of diagnosed mpox cases in LAC grew sharply, peaking in mid-August 2022 (Fig. 1A). By November of that year, cases had dropped rapidly, with only 31 cases being reported that month compared to 1033 in August. Since the start of 2023, mpox cases in LAC have been sporadic, mostly characterized by periods of low incidence followed by small clusters of infections usually found from May-July or December-January (Fig.1A) (19). Similar patterns can be seen in the number of third-generation mpox vaccinations administered whereby the majority of first and second doses were given in the summer and fall of 2022 followed by small increases in 2023 and 2024 surrounding early summer (Fig. 1C).

**Figure 1:**
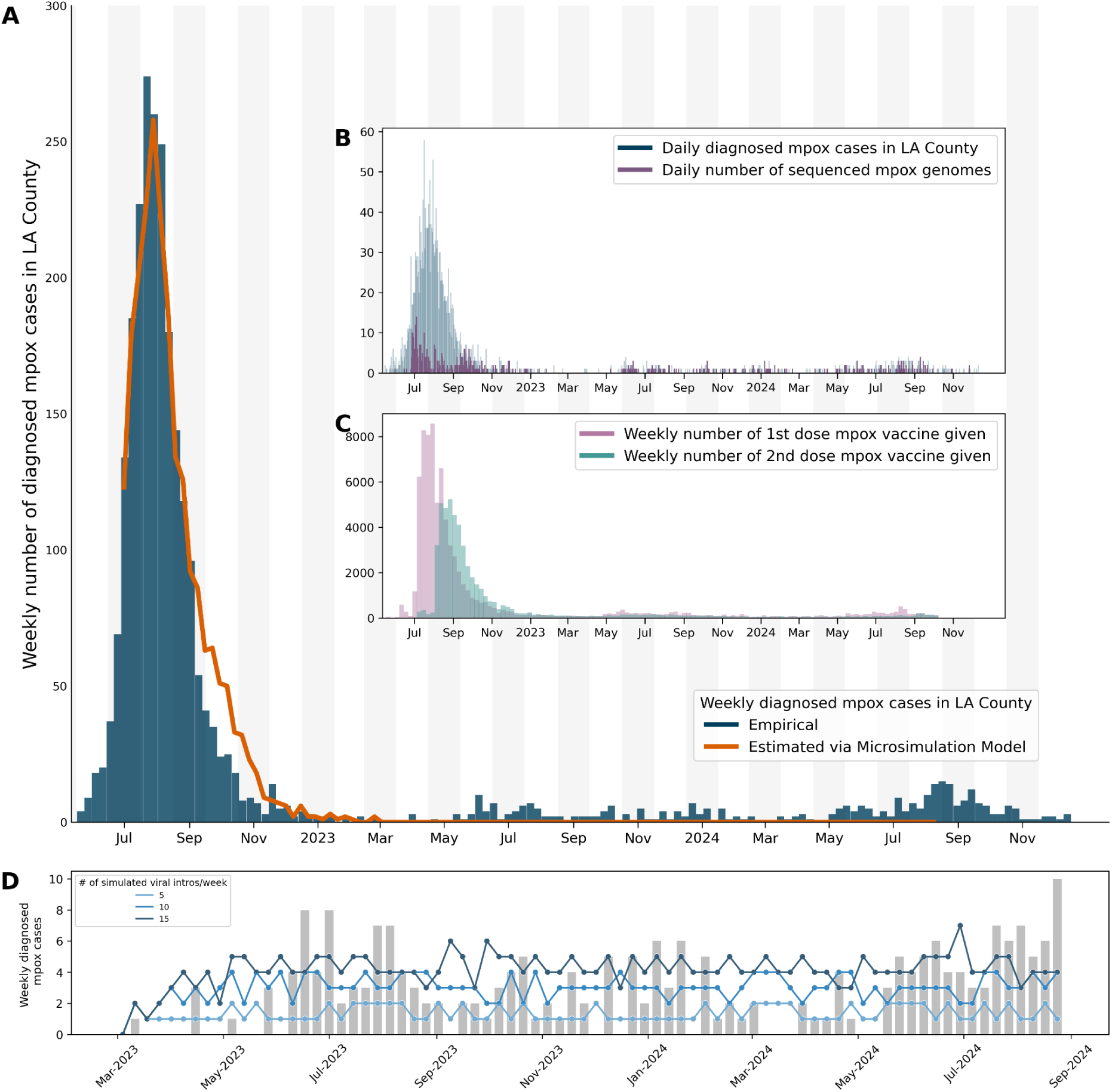
Mpox epidemiology and microsimulation modeling among men who have sex with men (MSM) in Los Angeles County. **(A)** Main figure shows the weekly number of diagnosed mpox cases in LAC from June 2022 through December 2024 (blue) with the number of diagnosed mpox cases simulated via our microsimulation model without viral importations overlaid in orange. Panel **B** shows the daily number of diagnosed mpox cases (blue) with the daily number of mpox sequences collected in LAC overlaid (purple). Panel **C** shows the weekly number of mpox vaccinations that were administered in Los Angeles County from June 2022 through October 2024 divided between the number of first doses (pink) and second doses (teal) given. **(D)** Scenario analysis of the impact of a constant force of viral introductions on our microsimulation model from March 2023 onwards. The empirical mpox case counts are represented by the gray bar chart while the simulated cases are represented by the blue point and line charts with an increasing number of viral importations per week.

In order to understand these low-incidence dynamics, we extended our previously-described microsimulation model of mpox tailored to the epidemiology and population structure of men who have sex with men (MSM) in LAC (18) to simulate the years 2023 and 2024. Specifically, our microsimulation model tracked mpox dynamics by age, race/ethnicity, and HIV status, and was calibrated and validated against LAC surveillance data (See *Methods*). While mpox affected more than just MSM (20,21), the an estimated 95% of mpox cases in the US have been among MSM (22), leading our model to be focused on this population.

Our model was able to accurately capture the number of diagnosed mpox cases in LAC through the beginning of 2023 (Fig. 1A, orange line). It showed, however, that without viral introductions into the county, mpox incidence would have been expected to drop to zero by March 2023, indicating that introductions are necessary for maintaining low mpox incidence (Fig. 1A). In order to test the impact of introductions in maintaining low, but not zero, incidence, we conducted a scenario analysis by simulating constant viral introductions every week into demographic strata randomly selected proportional to their population size. With the same model parameters calibrated after August 2022, the model required a large number of mpox introductions (> 5 per week) to maintain ongoing transmission similar to the empirical number of cases (Fig. 1B). This demonstrates that importations could be a key mechanism for maintaining ongoing, low-level prevalence.

### Periods of high viral introduction promote heavy-tailed transmission clusters that maintain low-level incidence

Estimating the empirical number of introductions into a region is difficult with case counts alone. Instead, we can leverage pathogen genomes to estimate the lower bound of the number of introductions. Since the start of the epidemic, the LAC Department of Public Health has sequenced a high volume of confirmed cases, leading to the number of sequences collected increasing as more cases were detected (Fig. 1B). While a low percentage of estimated cases were sequenced at the beginning of the 2022 epidemic, the majority of the months in 2023-2024 had more than 50% of the estimated mpox cases sequenced, allowing for local-scale phylodynamic investigation into ongoing local mpox transmission (Fig. S1).

To investigate transmission dynamics into LAC, we analyzed 497 mpox genomes sampled in LAC through December 12, 2024 alongside all available contextual sequences from around the world by creating a time-resolved phylogeny using Nextstrain (Fig. S2) (23). We also analyzed the inferred ancestral locations over time (Fig. 2) via discrete trait analysis focusing on the sequences from LAC (See *Methods*). The majority of LAC clusters in 2023-2024 were found to be part of lineage B.1.20 with one outbreak cluster consisting of lineage B.1.22 (Fig. 2A). While a large part of introductions into LAC in 2022 was inferred to come from global regions outside of North America, we found that in 2023-2024 introductions from within North America, primarily New York City and other parts of California, dominate (Fig. 2B). By inferring the location of viral exports from LAC, we found that, of the sequenced areas and viruses, about half of the viral exports from 2023-2024 were to other California regions, while the other half were mostly into Cook County, Illinois, and New York City.

**Figure 2:**
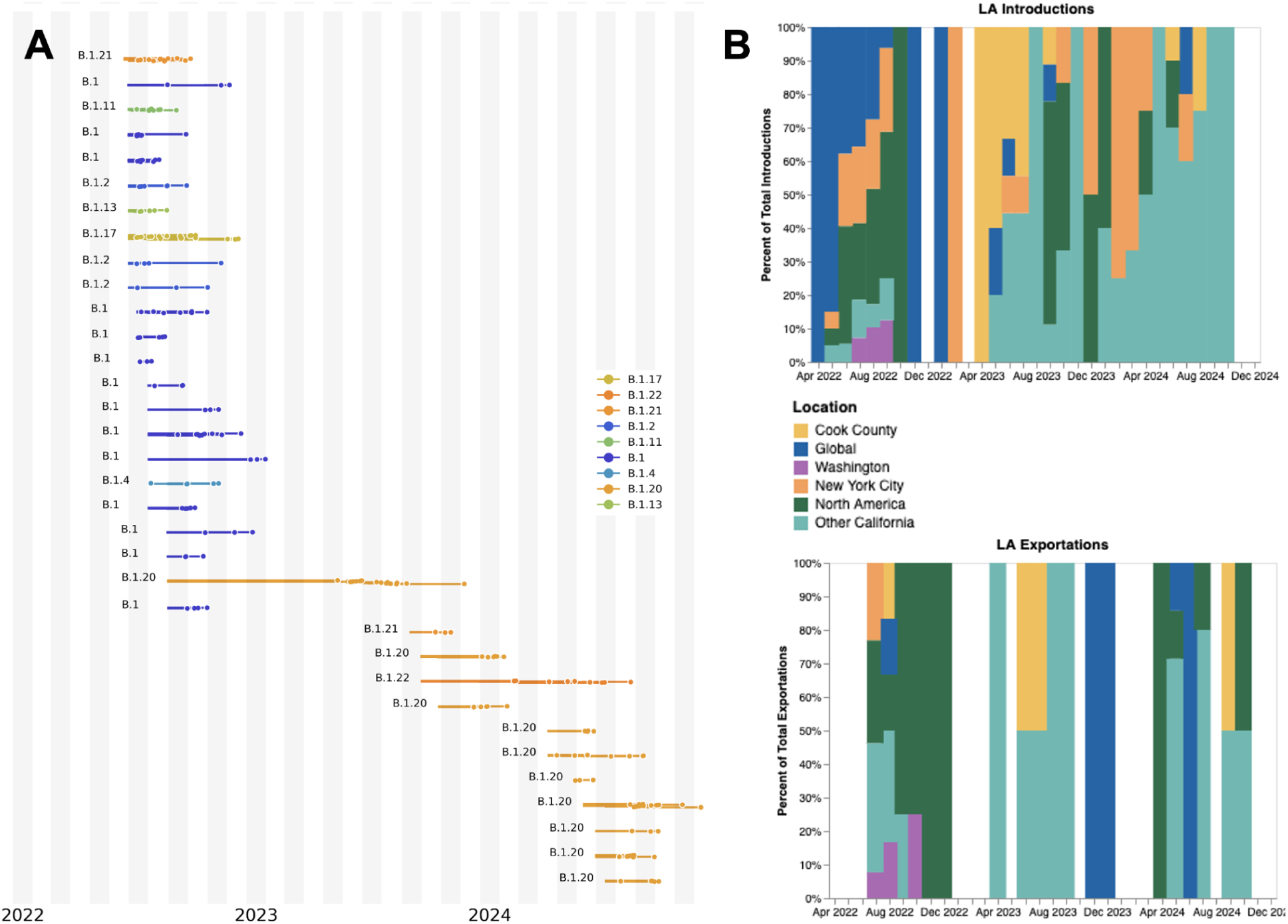
Genomic diversity, source of introduction, and location of exportation of mpox clusters in Los Angeles County. We analyzed more than 7,500 publicly available mpox clade IIb genomes from around the world via maximum likelihood phylogenetics using Nextstrain **(A)** Here, we show an exploded tree view of the maximum likelihood phylogeny that only includes the local outbreak clusters inferred to be in LAC via ancestral trait reconstruction (using Nextstrain’s augur traits functionality). Only clusters with more than three sequences are shown for clarity. The colors represent the assigned lineage of each cluster, showing the changes in mpox lineages circulating over time. **(B)** The plots on the right represent the inferred source of these imported clusters (top) as well as the location of viral exportations from LAC (bottom). The colors are shared between the two graphs and were constructed to focus on large metropolitan US cities and areas that have the highest level of mpox sequencing effort. The exportations and importations per month are normalized to 100% to highlight relative changes over time.

We then split the sequences into local outbreak clusters using parsimony-based clustering to identify groups of sequences whose ancestral states were inferred to be in LAC (see *Methods*, Fig. 3A). In total, we identified 287 clusters with the majority of them being of size 1 (n = 131). The distribution of identified outbreak cluster sizes is right-skewed, similar to the heavy-tailed sexual network distribution that was characteristic of the 2022 mpox epidemic where the majority of introductions resulted in no secondary infections and small number of introductions resulted in a large number of secondary cases (Fig. 3A top inset) (13). While we expected the total number of clusters identified to be affected by the sequencing both within and outside of LAC, we saw a very limited impact in our sample due to the high amount of sequencing worldwide and within LAC (Fig. S3).

**Figure 3:**
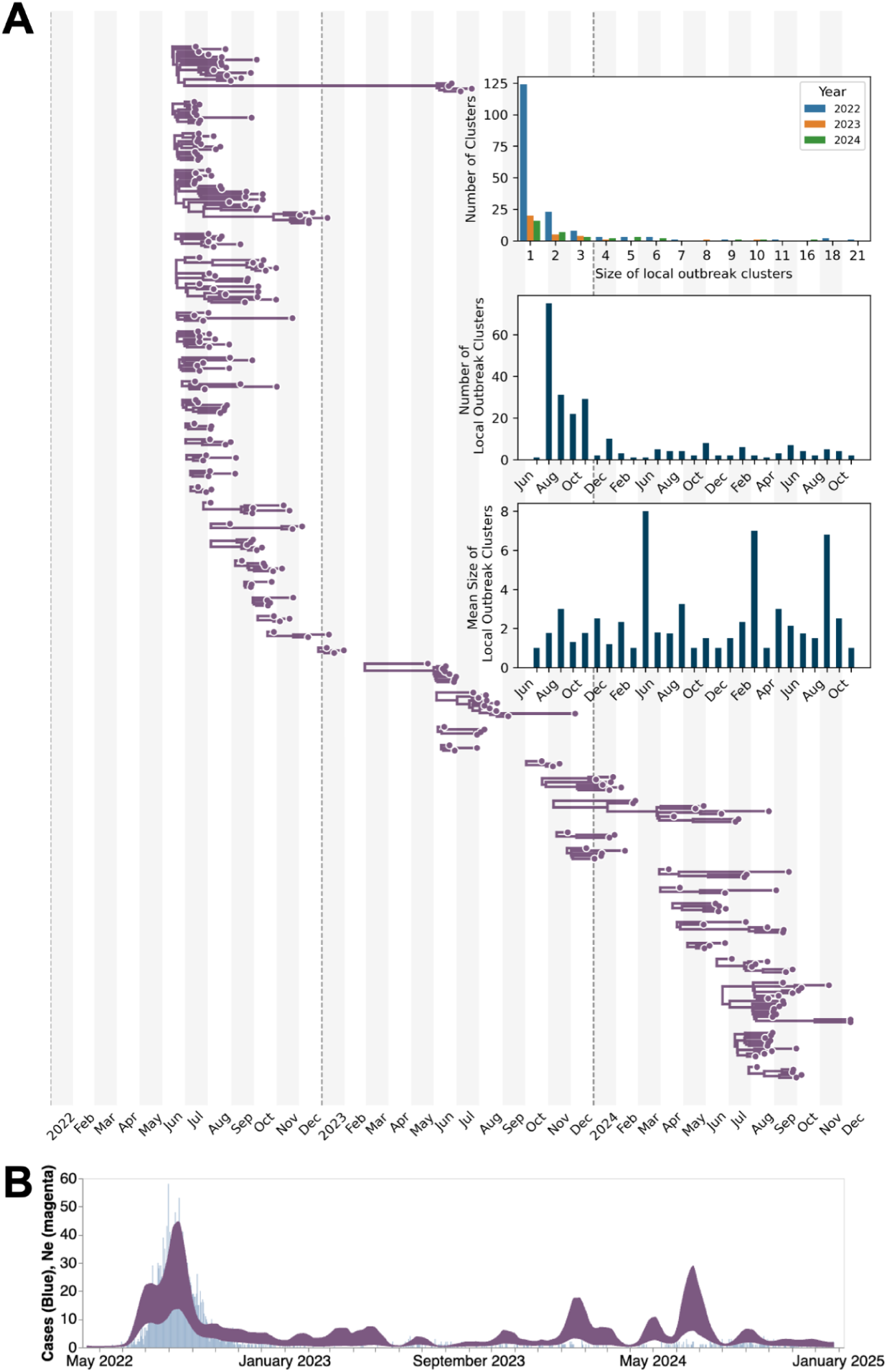
Local Los Angeles County dynamics estimated via Bayesian phylodynamics. **(A)** Maximum clade credibility (MCC) tree summary from local outbreak clusters of 497 sequences showing clusters with more than two sequences. **Top inset** represents the size distribution of the identified outbreak clusters by year; the **middle inset** is the number of identified outbreak clusters by month; and the **bottom inse**t represents the mean size of local outbreak clusters over time. The month is determined by the date of the earliest sequence in each cluster. **(B)** Estimates of effective population sizes (Neτ in years) from May 2022 through December 2024 (dark purple) plotted on top of the weekly number of diagnosed mpox cases (light blue). The coalescent time scale depends on both effective population size Ne (number of effective individuals) and on generation time τ (years per generation), resulting in Neτ being a measure of coalescent time scale in years.

We modeled the local mpox dynamics via a multi-tree coalescent phylodynamic approach conditioned on the *a prior* identified outbreak clusters (see *Methods*). In order to inform our estimates of transmission dynamics using both genomic and epidemiological data, we also developed a correlated case-based prior on the effective population size estimates using the weekly number of diagnosed cases smoothed using a 3-week moving average (see *Methods)*.

Our case-informed phylodynamic estimates of viral effective population size (*Ne)* were able to capture the temporal trends of empirical case data better than phylodynamic models informed by sequences alone (Fig. 3B, S4). We found time periods with higher *Ne* than expected by case counts alone, such as during the winter of 2023 or summer of 2024 where our *Ne* showed an increase in viral population size while case counts remained relatively constant, suggesting underdetected transmission (Fig. 3B).

Through our phylodynamic analysis, we were also able to estimate the date of importation for each identified LAC local transmission cluster, based on the most recent common ancestor time of each cluster, which provides a lower bound on the introduction time (Fig. 4A). The majority of introductions occurred during the summer of 2022, at the height of the 2022 mpox epidemic. In addition to this peak, we also saw the rate of viral introductions increase between February and June and August through October of each subsequent year (Fig. 4A-C).

**Figure 4:**
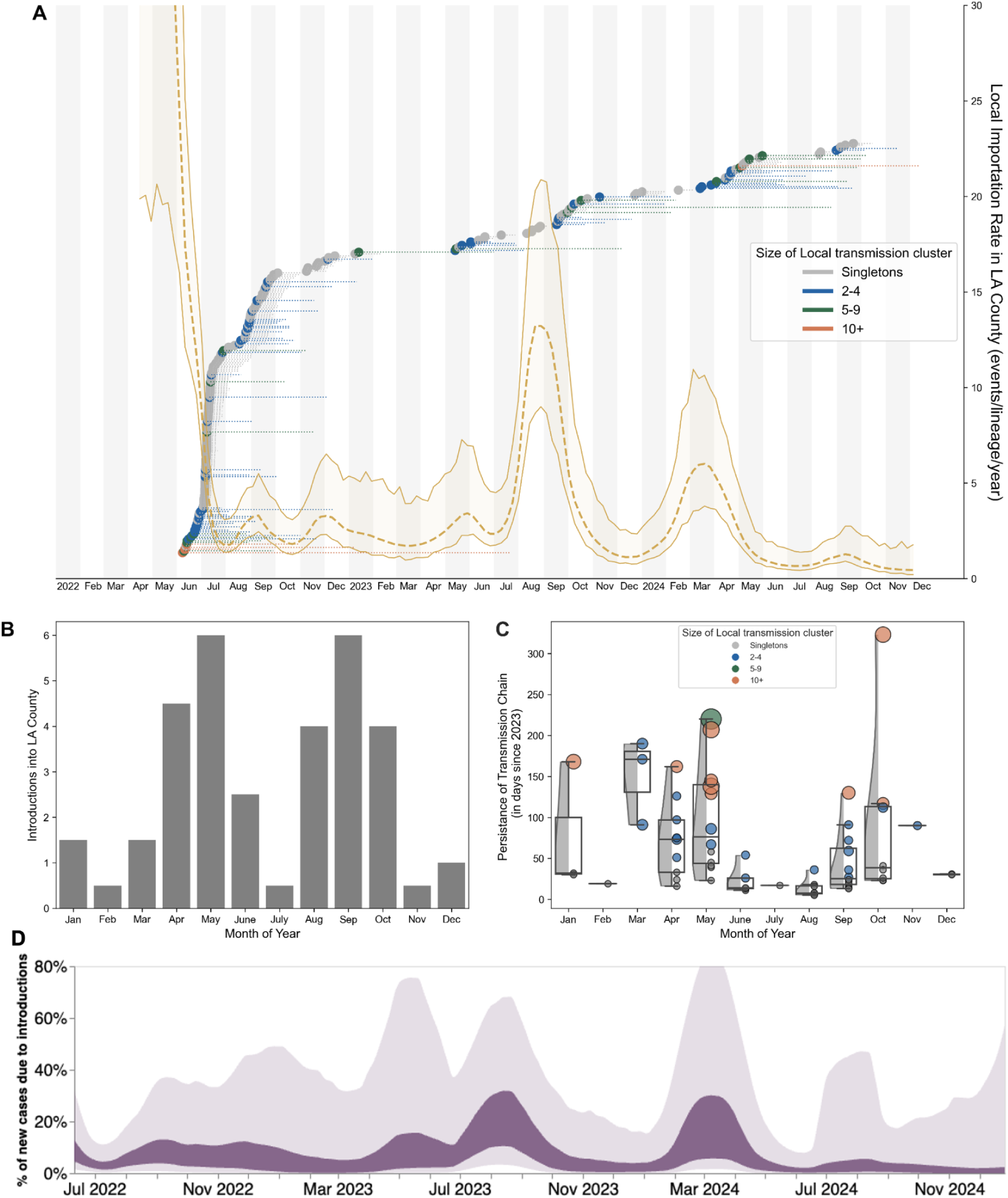
Patterns of viral introductions into Los Angeles County. **(A)** Here we plot the time of introduction for each local outbreak cluster estimated via our multitree coalescent approach, colored by the size of the resulting transmission cluster. The dashed line coming out of each point represents the time from the estimated date of introduction to the date of the last sequence sampled in the cluster (i.e. persistence). The yellow plot represents the time-varying rate of viral introductions estimated directly via the multitree coalescent, with the dashed line representing the median value and the upper and lower bounds representing the 95% highest posterior density (HPD). **(B)** Total number of viral importations into LAC per month since January 2023. Number of importations are adjusted by the number of observations in the sample. **(C)** The persistence times of downstream clusters by month of introduction since January 2023 with the boxplot plot representing the interquartile ranges, and the half violin plot representing the distribution of values. Scatter points are colored based on the size of the resulting transmission cluster. **(D)** The percentage of new cases due to introductions was estimated as the relative contribution of introductions to the overall number of infections in the region.

While the majority of introductions resulted in singletons (lead to only one sequenced genome), we found evidence of large transmission clusters introduced in both 2023 and 2024 during those months with a high force of introduction (Fig. 4B). Given the presence of these large transmission clusters, we subsequently estimated the persistence time of each cluster (estimated as the time, in days, between the inferred date of introduction and the sampling date of the latest sequence in the cluster). The persistence times for the largest clusters showed a maintenance of low-level case counts, often extending from one peak of introductions until the subsequent increase (Fig. S5A).

We found that unique introductions on average constitute around 20-30% of estimated new cases in LAC (Fig. 4D, dark bands). In periods of low case counts, we see a wide uncertainty (Fig. 4D, light bands), highlighting the potentially increased impact of introductions during those time periods. Of note, the phylodynamic estimation of the percentage of cases that are due to introductions only counts unique introductions and not secondary local spread after introduction. This formulation underestimates the impact of introductions on local case counts (See *Methods*) especially in light of how essential viral introductions were in the ability of our microsimulation model to recapitulate empirical trends (Fig 1A).

The rates of introduction and the percentage of cases from introductions were highest at the times closely prior to the two larger outbreaks in 2023 and 2024 (such as in March 2024), suggesting that outbreaks elsewhere and subsequent introductions in LAC were strong drivers of these outbreaks. When we explored the relative success of importations compared to ongoing lineages over two month time periods (Fig S6), we found further support of the high impact of introductions, especially in 2023, where introductions not only represented a large proportion of lineages in each time period (often more than 50%), but also were successful in creating downstream descendants that dominated local circulation in a short amount of time (Fig S6, green lines).

We used our effective population size estimates to calculate *Rt*, the time-varying effective reproductive number (Fig. S7A). We combined our estimates of *Rt* together with our quantifications of the percentage of new cases due to introductions to separate out the individual contributions of introductions and local transmission on *Rt.* We found that increases in *Rt* often follow increases in the percentage of cases due to introductions. While changes in the mean infectious period used to calculate the percentage of cases that are due to introductions and Rt (see *Methods)* impacted the variability and magnitude of our results, the patterns of interplay between introductions and local transmission remain the same (Fig. S7B-C). Comparison of *Rt* estimated from our phylodynamic analysis with *Rt* from empirical case counts alone for 2023-2024 showed similar dynamics when considering the combined impact of both importations and local transmission (Fig. S8B). Removing the influence of viral importations dropped the median *Rt* estimate close to 1 with high variability.

Additionally, given that the probability to observe a cluster of a given size is determined by the effective reproduction number *R* across a time period, transmission heterogeneity as estimated via the dispersion parameter *k*, and the fraction of infections sequenced (24,25), we explored how the probability to observe a cluster of size 16 (knowing we observed 64 clusters from 2023-2024) is impacted by *R* and *k* (Fig S8A), assuming that 5.5% of infections were sequenced (6). We find that for a value of *k* around 0.36, which is similar to what was estimated for previous mpox outbreaks and during the 2022 epidemic (6,26,27), it is highly probable to observe our max cluster size of 16 even with R values as low as 0.45, suggesting that the true R could be lower than 1. We estimated the reproduction number *R* from the distribution of sequenced cluster sizes (Fig 3A, top inset) from the same time period and found an *R* lower than one (Fig S8B), further suggesting that the true R for the time period is lower than one and that accounting for introductions can help partially correct the overestimation of local *Rt*.

We tested the ability of our approach to correctly estimate our parameters of interest via simulations (See *Phylodynamic Simulations* under *Methods)*. After simulating a local mpox outbreak with a constant force of introduction and superspreading with two different sequencing schemes (assuming all or 50% of cases sequenced), we found that our case-based prior approach is better at capturing temporal trends than analyses using sequences alone (Fig. S9-10). While the scenario with a Skygrowth prior on the growth rate analyzing sequences alone had the highest R^2^ value when comparing estimated *Rt* and the percentage due to introductions with simulated values, the 95% HPD intervals often failed to include the true value, while the Skyline prior of the *Ne* informed by case counts had a similarly high R^2^ while more often containing the true parameter value within the 95% HPD intervals. The case-informed Skyline prior was also found to be more robust to having 50% fewer genomes available when compared to the same model with a Skyline prior but without any case information. Ultimately, all three specifications of our model are able to capture the simulated dynamics, showing the utility of genomic information to inform investigations into local mpox dynamics as well as the added benefit of incorporating epidemiological information into our phylodynamic analyses. Our phylodynamic results are robust to differences in substitution model specification (Fig. S4).

### Combining phylodynamics and epidemiological microsimulation suggests a return to baseline sexual behavior in 2023-2024

From our phylodynamic results, we estimated the absolute number of viral importations into LAC over time (Fig. 4B and S6B). This allowed us to reparameterize our microsimulation model to incorporate an estimated force of introduction.

Briefly, our microsimulation model included a dimensionless calibration parameter, here referred to as the Infectivity Scalar (α), which we vary over time (See *Microsimulation Model,* under *Methods)*. The Infectivity Scalar was designed to modify the transmission impact of infected individuals on the susceptible population within their respective demographic group. Given that our model accounts for assortative mixing patterns between demographic groups as well as for the development and waning of vaccination-induced immunity, the Infectivity Scalar largely serves to capture changes in behavior throughout time, representing the relative risk of disease spread.

For the first five weeks following the surge of mpox in LAC in 2022 (from early July to early August), α was calibrated to 2.2, establishing a baseline for the impact of sexual behavior on mpox transmission (See (18), visually represented in Fig. S11). Following those first five weeks, α was lowered to 0.7 to align model outputs with empirically observed case estimates, representing a significant reduction in the risk of disease spread via changes in sexual behavior, in concordance with previously-documented reports (28,29).

After adding the time-varying weekly number of estimated introductions into our model, we found that our model is able to recapitulate a similar number of diagnosed mpox cases in LAC as in the empirical data (Figs 5A-B). To do so, however, required increasing α starting in March 2023. We tested different α levels from 0.7 to 2.2, whereby 0.7 represents the decreased sexual activity following the peak of the 2022 mpox outbreak in LAC, and 2.2 represents the baseline α during the beginning of the 2022 outbreak. By comparing the simulated number of mpox diagnoses with the empirical case counts from LAC, we found the optimal α to be 2.0, which represents a significant return in sexual behavior when compared to late 2022 (Fig. 5A-B, dark blue lines). Given that phylogenies only capture successful introductions and that only a number of these introductions are sequenced, the phylodynamic-estimated number of introductions into LAC is expected to be a lower bound. We found that even when doubling the number of estimated importations, both time-varying viral introductions and a subsequent increase in sexual behaviors are still needed to recapitulate the empirical number of mpox cases, adding further support to our conclusion (Fig. S12). We also tested the sensitivity of our findings by increasing and decreasing the average effectiveness of mpox vaccination against infection after one year by 25% and find that the cumulative number of empirical mpox cases still falls within the 95% CI of α of 2.0 (Fig S14).

**Figure 5:**
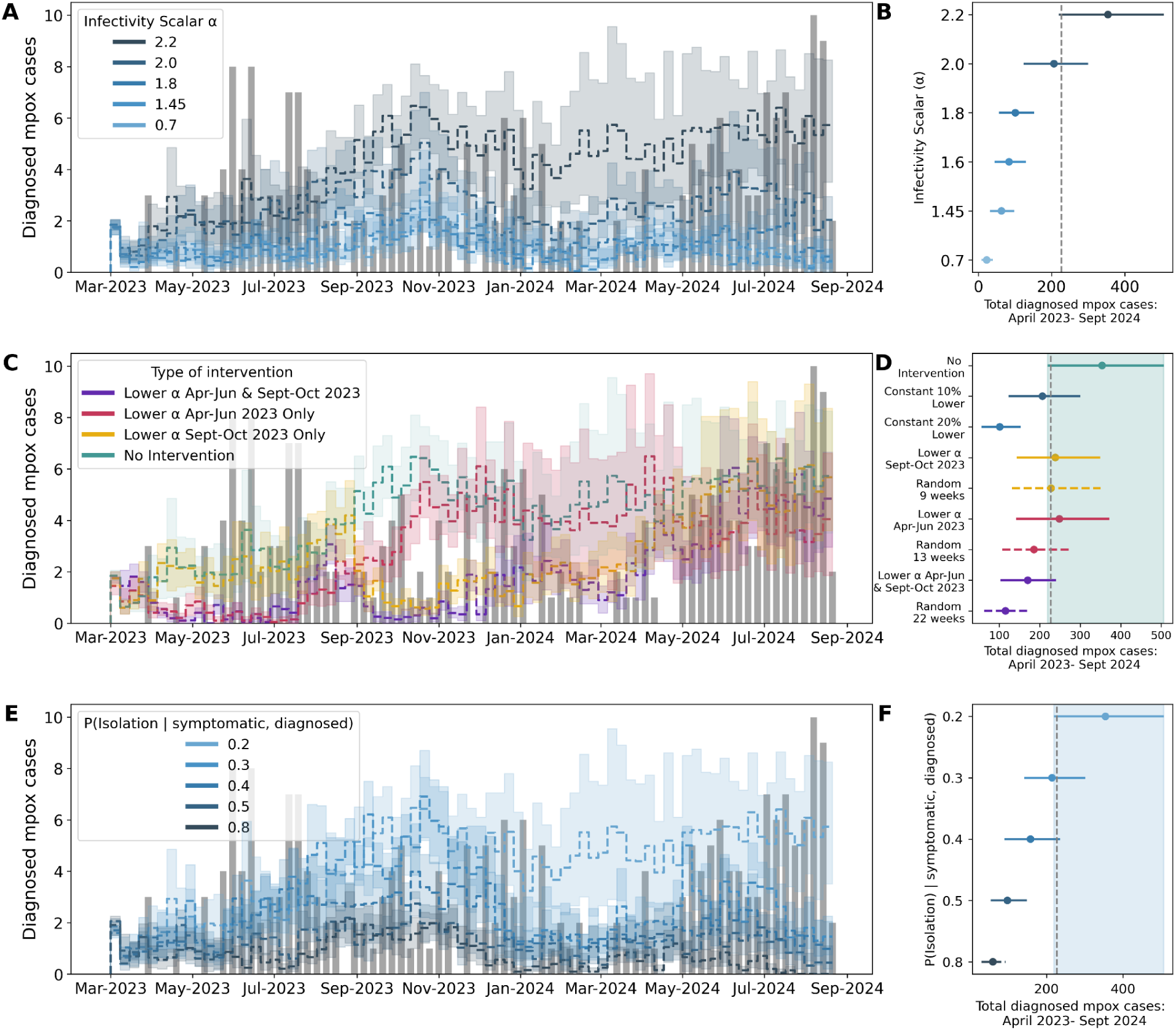
Factors maintaining mpox prevalence and modeling counterfactual public health interventions. After parameterizing our microsimulation model with the number of viral importations estimated via phylodynamics, **(A, B)** we explored the α that best explains the empirical weekly number of diagnosed mpox cases (gray bars). Line graphs represent the mean weekly number of mpox diagnoses simulated using increasing α. Given the non-constant pattern of viral introductions seen in the phylodynamic analysis, we tested different counterfactual scenarios of public health interventions during specific time periods **(C, D)** represented by lowering the α to 0.7 while keeping the α at 2.2 during the remaining time. The bold yellow, red, and purple lines represent the simulated weekly number of diagnosed mpox cases under phylodynamic-informed interventions. In **D,** we also compared the impact of lowering the α for the same random number of weeks as each specified intervention as well as the impact of constantly lowering the α throughout the entire time period by about 10% and 20% to simulate a constant low-effectivity intervention. The green area represents the upper and lower bounds of the “No Intervention” scenario. We also tested the effect of increasing the probability of isolating upon a symptomatic individual receiving a positive mpox diagnosis on the simulated number of diagnosed mpox cases **(E, F).** In **F,** the light blue area represents the bounds of the base model scenario with an α of 2.2. In **A, C, E**, the grey bars represent the empirical number of mpox diagnoses in LAC. We calculate the uncertainty of our microsimulation results via bootstrapping to estimate 95% uncertainty intervals for each weekly simulated estimate. For **B, D, and F**, the dashed line represents the total empirical number of diagnosed mpox cases from April 2023 through September 2024.

We also calculated *Rt* from our microsimulation model by tracking the weekly number of secondary cases per each infectious individual (Fig S8B, S15). For α of 2.0, we found a median *Rt* to be 0.53, which is lower than the median *Rt* estimated from other methodologies but in concordance with the *R0* estimation from cluster distributions (Fig S15). Additionally, we see a median *Rt* significantly below 1 even at a higher α of 2.2 (median *Rt:* 0. 62, Fig S15), which in Fig 5A-B resulted in an overestimate of the estimated number of mpox cases. This finding, together with the *R* calculated from the distribution of cluster sizes, suggests that the true median *Rt* during 2023-2024 is significantly below one.

### Counterfactual scenario modeling reveals the potential impact of public health interventions in curbing mpox case counts

We employed our phylo-informed microsimulation model to explore the impact of various potential public health interventions on mpox spread in LAC from 2023 through 2024. Given the time-varying nature of viral introductions seen in the phylodynamic analysis (Fig. 4), we tested the impact of uniformly lowering transmission pressure during the months of highest viral introductions (April through June and September through October in 2023) by lowering the α to 0.7 (Fig. 5C-D). We lower α to simulate significantly reduced sexual behavior prompted via an unspecified public health intervention. During the months without public health intervention, we kept α at the baseline of 2.2. We tested the specificity of our proposed counterfactuals by lowering α to 0.7 for the same number of weeks but selected at random (Fig. 5D, S13). Our analyses showed that while lowering α during one period of high viral introductions resulted in lower but albeit similar cumulative numbers of total simulated cases when compared to the scenario without intervention, targeting both periods of high-intensity introductions during 2023 resulted in an average reduction of almost half of all simulated cases over the entire time period. (Fig. 5C-D, dark purple line). In Fig. 5D, we also compared the effect of high-intensity public health interventions only during the high importation months vs having a low-efficacy intervention occurring throughout the entire time period. We simulate this counterfactual by lowering α by approximately 10% and 20% (α of 2 and 1.8 compared to 2.2, respectively). We find that intensely targeting just one time period during 2023 had a similar impact as a constant low efficacy intervention that lowered α by 10% for the entire time period between 2023-2024. We also found that intensely targeting both time periods of high viral importation volume just in 2023 had a similar effect on average in controlling the numbers of estimated total mpox cases than did having a constant intervention that was only about 20% effective throughout the entire time period. Notably, our targeted counterfactual intervention strategies were applied only in 2023. Despite having significant reductions in simulated cases during that year, we see that by summer of 2024 our model simulates nearly identical weekly case counts for all counterfactuals when compared to our scenario without interventions (Fig 5C), highlighting how new viral importations, rather than continued local transmission, could be the main contributor of empirical case counts in LAC. The rebound in cases driven by viral importations in 2024 could explain the similar cumulative impact of our targeted interventions only in 2023 when compared to the same number of weeks selected at random (Fig 5D horizontal dashed lines, Fig S13).

We also analyzed the impact of increasing the probability of isolation given an infected individual was symptomatic and subsequently diagnosed with mpox (Figs 5E-F). To do so, we fixed α to the baseline of 2.2 and increased the probability of isolation starting at 0.2 which represents the baseline model. We found that increasing the probability of isolation by increments of 0.1 resulted in large decreases in the total number of diagnosed mpox cases throughout the time period.

## Discussion

After decades as a predominantly regional infection, the mpox virus spread globally in 2022, mostly via queer sexual networks. While the epidemic in 2022 has been extensively studied (5,6,15,16), very few studies have investigated the dynamics of mpox clade IIb in 2023-2024, when cases remain low and sporadic, but resist elimination (19). Here, we combine phylodynamic and microsimulation modeling approaches to describe the 2023-2024 dynamics of mpox transmission in Los Angeles County, a diverse, metropolitan US County with a high level of rapid genomic mpox surveillance. We show both the impact of imported mpox cases and the heavy-tailed pattern of local transmission define the sporadic nature of mpox cases in this large population center and how the return of typical sexual behaviors alongside a median *Rt* below one might explain the current case trends.

A major strength of our study is the combination of Bayesian phylodynamics inference and microsimulation modeling to help address these knowledge gaps. Both phylodynamic analysis and mathematical modeling have played a crucial role in understanding infectious disease dynamics as well as in informing public health decision-making (30). Independently, however, both methods have limitations: understanding the interplay of local transmission and viral importations is difficult via case counts alone (11,31), limiting the power of the microsimulation to capture these dynamics; phylodynamics works to understand shared ancestry as it relates to transmission, making it difficult to simulate counterfactual scenarios and capture dynamics of low-incidence pathogens. Prior work has helped highlight the utility of combining these two approaches (32,33) but has been limited by the use of deterministic compartmental models and maximum-likelihood phylogenetic methods that are sensitive to differential sampling.

Here, we jointly model both the rate of importation into LAC and local mpox dynamics using Bayesian coalescent phylodynamics, and use those results to inform a stochastic microsimulation model to understand low-incidence pathogen transmission and simulate counterfactual public health interventions. Our work is tailored to the local, heterogeneous demographic and epidemiological landscape of LAC and models the interplay between local transmission and introductions to understand local mpox dynamics. Ultimately, our study serves as a model for understanding factors that maintain low-level viral disease prevalence in a large, diverse, heterogeneous metropolitan US region.

Our analysis demonstrates that both local transmission and mpox importation contribute to the ongoing sporadic pattern of mpox spread in a large urban center. In alignment with other studies (13,15,34), we show that most mpox importations lead to a singleton (one case without onward transmission), but a small number of importations transmit to more than ten downstream cases, which are successful in dominating local circulation (Fig S6). This pattern suggests that identification and intervention, either via vaccination or behavior change, in the small proportion of mpox importations that lead to a large number of local cases could have an outsized impact on the overall dynamics of mpox spread. Local public health efforts to promote vaccination among communities disproportionately impacted by mpox that are community-centered and located in community settings through the use of mobile vaccination teams have encouraged vaccine uptake and provided education regarding sexual behavior harm reduction strategies to prevent mpox transmission (35,36).

Our results further show that importations of mpox in Los Angeles County varied over time, with a large number of importations occurring in mid-fall and -spring in 2023 and 2024. These insights may be particularly useful for the formulation and deployment of public health campaigns that promote vaccination and sexual behavior harm reduction strategies. Our counterfactual modeling showed that targeting both of these time periods has the potential to significantly reduce the number of mpox cases (Fig. 5B), allowing for more precise targeting of public health resources. Our results showed, however, that this form of public health action is most effective when both time periods are targeted, and that case counts will eventually return to baseline without recurring interventions, suggesting the need for periodic implementation of public health action rather than just singular, one-off interventions. When we compared our targeted interventions to a low-efficacy constant public health intervention, we found that high-intensity targeting during just 2-5 months when introductions were highest had a similar effect to continual, low efficacy interventions, suggesting that identification of high introduction time periods could allow for prioritization of public health effort and showing the high-impact potential of genomics and microsimulation-informed campaigns. Timing social marketing campaigns (37,38) and vaccine clinics based on patterns of mpox seasonality are promising, as are strategies that focus on raising awareness for travelers and their sexual networks (12).

Given that the estimates from both the distribution of cluster sizes and the phylodynamics-informed microsimulation model show a mean *Rt* below one (Fig S8), our results suggest that the true median *Rt* of mpox in LAC during this time period is most likely well below one (which is highly probable as seen in Fig S8A). This is in concordance with the observation that our microsimulation model needs viral importations to maintain a low case incidence following the 2022 epidemic (Fig 1D) and the similar case counts by the summer of 2024 in all our counterfactual scenarios in Fig 5C, suggesting that weekly case counts are driven by patterns of viral introductions more than by prior levels of local transmission.

The *Rt* results suggest a “stuttering chains” dynamic whereby viral importations result in a heterogeneous distribution of secondary cases but eventually go extinct, which is what we observed in Fig. 4A. Therefore, a consistent inflow of viral introductions is needed to maintain the low case counts. Due to the limitations of passive public health surveillance, stuttering chains can often become “entangled”, resulting in persistent case counts that result in an overestimation of the effective reproductive number as previously shown (27). This phenomena can be seen in Fig S5A where the majority of clusters go extinct quickly, but some overlap, giving the impression of constant incidence without exponential growth. Our “entangled stuttering chain” hypothesis is further supported by Figure S15, where our genomics-informed microsimulation model shows that even at an α of 2.2, which we know overestimates the number of diagnosed mpox cases in LAC (Fig 5B), the median *Rt* is still significantly below 1 (median *Rt* with α of 2.2: 0. 62), suggesting the true median *Rt* to be around 0.53 (with α of 2.0).

We expect our estimates of *R* from the distribution of cluster sizes to be artificially elevated as sequencing and phylogenetics is more likely to capture successful and larger clusters than introductions with no secondary cases, artificially increasing the mean cluster size and resulting in an overestimation of *R* (27). We note that there is high variability in the estimates of median *Rt* regardless of the methodology used (Fig S8), highlighting the difficulty of estimating these epidemiological parameters at periods with low incidence (39) and that methods that fail to account for the impact of introductions overestimate the *Rt* (31) (Fig S8). Reductions in disease prevalence are often not easily captured in coalescent models, as effective population sizes decrease linearly with decreasing prevalence, but also increase linearly with decreasing incidence (40,41). This limitation of coalescent models most likely explains why the average phylo Rt even when corrected for absolute number of introductions is not further below one, and highlights the importance of combining phylodynamics with microsimulation modeling for understanding pathogens of low-incidence. Together, our evidence suggests that the true median *Rt* of mpox in LAC from 2023-2024 is around 0.53 and that time-varying peaks of importations often lead mpox to establish stuttering chains in a densely-connected sexual network that can last until the next peak of introductions.

We used our phylo-informed microsimulation model to uncover factors maintaining the observed low-level mpox prevalence and to test actionable public health interventions. Since our microsimulation model, despite being informed with viral importation estimates, required a recalibration of the α parameter, our modeling suggested the low-level, but persistent number of mpox cases in LAC can be explained by a combination of waves of viral introductions, a median *Rt* below one, and a return to near-baseline sexual behavior in 2023-2024. Previous work using online surveys of MSM in North America have shown that more than 78.4% of surveyed individuals who had modified their sexual behavior in response to the 2022 epidemic had reversed their adaptations by May of 2023, showing the plausibility of our results (29). Of note, both sexual behavior and travel vary by season (6,42,43), often peaking in summer months with large LGBTQ+ events, when the 2022 outbreak in LAC began; therefore the baseline α of 2.2 may represent an upper bound of sexual activity since it was established using only five weeks between June-July 2022 (18). While our results offer a mechanistic explanation for present-day transmission dynamics and reveal potential avenues for public health interventions, other factors such as heterogeneity in immunity duration post vaccination or infection (44,45), or turnover of susceptibles potentially from younger individuals reaching sexual primacy might still be impactful. Future work that combines line-level metadata regarding each infection that contains information regarding age, infection history, vaccination status, and zip code of residence and is matched to viral genomic information could further elucidate the nuanced mechanisms promoting mpox transmission.

Given the potential return of baseline sexual behavior, the infection control strategies during the ongoing mpox outbreaks might be different than those during the 2022 epidemic (12,26,29). For example, we tested the impact of increasing the probability of isolation after a symptomatic, infected individual receives a positive diagnosis. We found that increasing the probability from 0.2 to 0.3 resulted in a lower number of diagnosed mpox cases than seen in empirical case counts, highlighting a potential target for public health intervention. Prior modeling work that accounts for the length of viral shedding has shown that isolating three additional days after mpox lesion resolution is sufficient to eliminate more than 95% of post-diagnosis transmission (46). The authors of that work also note that individual viral shedding kinetics are heterogeneous and that a testing-based isolation strategy could reduce the total time of isolation. Researchers have found, however, that individuals who have previously experienced mpox-like symptoms show a lower willingness to self-isolate after a positive diagnosis, suggesting the need for a more tailored approach for previously-infected individuals (47). Further work is needed to determine the most effective method of isolation that balances the risk of transmission with the desire for social and sexual contact. For example, prior research has shown that, after adjusting for relevant covariates, engaging in condomless receptive anal sex with an individual with mpox symptoms had the highest association with increased risk of mpox transmission (48), suggesting that a modification of sexual behavior rather than complete abstention could be a potential harm-reduction strategy. The authors found a potential association between sharing bedding or clothing and the risk of transmission in an unadjusted analysis but the association was lowered toward the null and nonsignificant when adjusted for relevant covariates, highlighting the need for further work on the risk of non-intimate contact in mpox transmission.

Our results have limitations to note. First, despite our use of all publicly available mpox genomes from LAC, the changing proportion of cases successfully sequenced and uploaded from LAC (Fig. S1) will impact the chance that a case shows up in our data through the period studied. Our phylodynamic analyses are conditioned on the inferred sequence clusters from LAC which are dependent on the integration of contextual sequences from US and global regions into a temporally-resolved phylogeny. It is possible that differential sampling from other locations could impact our identified clusters, and ultimately our estimates on the rate of introduction. Our simulation analysis where we downsample different proportions of contextual sequences, however, shows a limited impact on the number of clusters identified as well as the mean cluster size (Fig. S3A). Limited mpox sequence diversity, especially during periods of rapid transmission such as at the beginning of the 2022 epidemic, could affect our ability to break up larger clusters. This might lead to collapsing multiple introductions into LAC into shared clusters, although prior work has shown that APOBEC3 editing associated with human-to-human transmission of mpox results in a mutation rate similar to RNA viruses (6,49,50). While it would be optimal to explicitly account for locations outside of LAC, ideally through a GLM approach that would also help ameliorate the limited sequence diversity, prior work has shown the high computational cost of these approaches (6). Our approach allows for Bayesian analysis of mpox dynamics within LAC in less than a day, while phylodynamic approaches with a GLM and explicit modeling of different contextual locations have been shown to take upwards of a month. Bayesian coalescent models assume random sampling of infected individuals, meaning that targeted sampling, such as superspreader events or contact tracing, could bias our phylodynamic estimations, although our simulation results show that our models are able to robustly capture complex simulated dynamics that incorporate superspreading (Fig. S9-10). Additionally, phylogenies only capture successful introductions into LAC that were ultimately sequenced, meaning that parameterizing our model with the estimated absolute number of introductions inherently underestimates the number of true viral introductions. While informing our model with the estimated absolute number of introductions was necessary due to the underlying microsimulation model structure, our main conclusion – both time-varying viral introductions and an increase to near-baseline sexual activity are needed to explain current mpox dynamics in LAC – is robust, even when we doubled the number of importations estimated (Fig. S12). Future work should focus on parameterizing models with the rate of introductions or the percentage of cases due to introductions.

While we calibrated our microsimulation model using vaccination data from the LAC Department of Public Health (51), the model does not explicitly account for seasonal variations in mpox vaccination rates, such as the observed increases from May to September 2023 and from July to September 2024 (Fig. 1C). Despite this, the microsimulation model successfully captures the overall vaccination trends by dosage and subgroups, including HIV status, as illustrated in Figure S17. Given the low likelihood of reinfection following mpox infection (52), our model assumes only waning vaccine-induced immunity, which may increase the number of susceptible individuals over time. Of note, the Infectivity Scalar (α) is a global parameter and does not capture heterogeneity in sexual behaviors or other unmeasured mechanisms that might modify the risk of transmission. While the model incorporates age- and race-stratified mixing patterns, individual-level transmission risk variation within those demographic groups is not accounted for, and nor is variation over time within those groups, although prior work has shown that collective behavior assumptions can effectively approximate population-level mpox dynamics (52). Additionally, changes in α in our model were derived through calibration and are not based on direct observation; as such, an unobserved, time-varying effect that modified transmission rates during the analysis period could lead to bias in our α calibration. To mitigate this possibility, we account for as many known modifiers of mpox incidence as possible given the available data (the model includes testing, diagnosis, treatment, disease progression, and recovery rates; see (18) for details). Additionally, our counterfactual scenarios simulate only a generalized increase in α during specified periods, which may not fully reflect the true dynamics of disease interventions. Finally, while we addressed stochastic uncertainty using multiple model iterations and bootstrap resampling, we did not perform a full probabilistic sensitivity analysis across all model parameters. However, we partially assessed parameter uncertainty by conducting sensitivity analyses on key inputs, such as vaccine efficacy.

In conclusion, our results suggest that the persistent transmission of mpox in 2023-2024 in a large urban US county can be explained by a combination of time-varying viral importations, a median *Rt* significantly below one, and the return of baseline sexual behaviors that were altered during the 2022 mpox epidemic. Our modeling supports that education and support for mpox patients such that they can maintain isolation from sexual networks while infectious and symptomatic may decrease the number of mpox cases in large urban centers. Further, messaging and targeted vaccination around travel, especially in mid-fall and -spring, may decrease the number of clusters generated by mpox importations during this time. Our synergistic phylodynamic and microsimulation approach can reveal factors in ongoing mpox dynamics that lead to significant local spread and can be leveraged by local health departments for specific health interventions.

## Methods

### Mpox cases counts data source

Data on the number of diagnosed mpox cases in Los Angeles County were downloaded from the Los Angeles County mpox data dashboard (http://publichealth.lacounty.gov/media/monkeypox/data/index.htm/ ; last accessed on 01-20-2025).

### Estimation of mpox incidence, prevalence, and effective reproduction number via case counts

To jointly estimate mpox case incidence, prevalence, and effective reproduction number, we used the renewal equation framework from Figgins and Bedford (53). Similar to Paredes et al (6), the time-varying effective reproduction number (i.e. the average number of secondary cases infected by a single primary case) was modeled using a 4th order spline with 5 evenly spaced knots assuming a discretized gamma-distributed generation time with mean 12.6 days and standard deviation 5.7 days (54). Case counts were modeled using a Poisson distribution. This model produces posterior estimates of daily incidence (defined as the number of newly infected individuals in absolute counts) and effective reproduction number.

Models were fit to aggregated case counts for each region using full-rank stochastic variational inference. Optimization was performed using the ADAM optimizer with learning rate 4e-3 and for 50,000 iterations and 500 samples were drawn from the approximate posterior.

As an additional comparison, we also independently estimate *Rt* using case counts alone via EpiFilter, which has been found to be more robust during periods of low case incidence (39). To calculate the Rt, we assume a gamma-distributed serial interval of 8.7 days estimated by Ponce et al (55).

To estimate the proportion of cases that were sequenced, mpox incidence estimated by the above renewal equation framework was aggregated into monthly estimates for year; the same was done for the number of sequences from LAC. The monthly incidence was then divided by the number of monthly LAC sequences. Due to the limitations of the renewal equation framework (not accounting for overdispersion, strong smoothing) as well as the stochastic nature of genomic sequencing, some months were found to have more sequences than estimated cases. In this situation, we created a ceiling of 100% as a way to bound the variance of estimates.

### Microsimulation Model

#### Model Overview

In this study, we utilized an individual-based Markovian microsimulation with a weekly cycle to project the dynamics of the 2022 mpox outbreak among MSM in LAC (18). The model simulates transmission, diagnosis, vaccination, isolation, and treatment, with individuals transitioning through health states (susceptible, exposed, asymptomatic, symptomatic, diagnosed, isolated, treated, and recovered) based on probabilities that vary by age, race/ethnicity, and HIV status (see model schematic in Fig. S16).

The model includes both preventive vaccination for susceptible individuals and post-exposure prophylaxis to reduce the likelihood of symptom development after exposure. Symptomatic individuals who are aware of their infection may receive treatment and be isolated to reduce further transmission. In the model, mpox transmission is modeled to occur exclusively through sexual contact during the symptomatic phase, using a sexual mixing matrix capturing partnership patterns and transmission risks across age and race/ethnicity (see Appendix Figure 3 in (56)). The model also accounts for the higher susceptibility to mpox among people with HIV. New susceptible individuals enter the model each week at age 15, while exits occur due to non-mpox-related deaths. Parameter values can be found in Liang et al. (18).

The model was originally calibrated and validated against data from July 2022 to March 2023, including diagnosed cases and vaccination uptake by age, race/ethnicity, and HIV status, as detailed in Liang et al. (18). Building on this foundation, we refined the model for the current analysis by incorporating updated vaccination data and vaccine efficacy assumptions, while keeping all other model parameters consistent with the original model unless otherwise noted.

Specifically, we incorporated updated vaccination uptake disaggregated by dosage and HIV status, covering the period from March 2023 to October 2024 (see Fig. S17). We also adjusted the model to reflect the potential waning vaccine efficacy, assuming a linear decline to 50% of its initial effectiveness one year after vaccination (44,45,57). The enhanced model was first run from July 2022 to March 2023 to generate a realistic baseline population, which was then used to simulate disease dynamics over an 85-week period, spanning from March 12, 2023, to October 27, 2024.

#### Modeling Infection Risk

The probability of mpox infection for an individual, denoted as *P*(*infection*), was determined by the interplay of multiple factors capturing demographic variation and behavioral dynamics in the population. In our weekly-cycle model, *P*(*infection*) represents the probability that a susceptible individual becomes infected within a single week. Specifically, it was defined as:

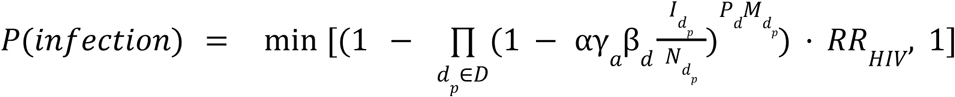

Where:

*D*: Set of demographic groups considered in the mixing matrix
*d*_*p*_: Partner demographic group
α: Infectivity scalar
γ_*α*_: Age-specific calibration parameter for the susceptible individual, where
*α* ∈ {15 − 24, 25 − 34, 35 − 44, 45 − 100}
β_*d*_: Race/ethnicity-specific calibration parameter for the susceptible individual, where
*d* ∈ {*Black, Hispanic, White*}
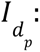: Number of infectious (not isolated) individuals in partner demographic group *d*_*p*_
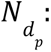: Total number of individuals in partner demographic group *d*_*p*_
*P*_*d*_: Average number of partners for an individual in the demographic group *d*
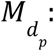: Probability that a susceptible individual mixes with partners in demographic group *d*_*p*_
*RR*_*HIV*_: Relative risk multiplier applied to individuals with HIV to reflect increased susceptibility

This formulation captures the combined effects of group-specific transmission risk, partner mixing patterns, and variation in partner counts, allowing infection risk to vary across age, race/ethnicity, and sexual network structure.

#### Calibrating and Estimating the Infectivity Scalar (α)

The microsimulation uses a calibration parameter we refer to as the ’Infectivity Scalar’ (α ) to adjust the probability of infection and fine-tune transmission dynamics. In the initial model setup, α was calibrated through a grid search across a range of potential values and set to 2.2. This value was chosen based on its alignment with the empirical trends observed during the early phase of the mpox outbreak in LAC, indicating high transmission levels prior to August 2022. Subsequently, α was recalibrated to 0.7 in response to a noticeable decline in mpox cases. This adjustment likely reflects the impact of enhanced public health guidelines and changes in public behaviors and allowed the model to effectively capture the decline in transmission risk.

The calibrated model accurately reflected the observed transmission trends. By December 2022, the seven-day average number of new cases had dropped below two, and by March 2023, the model predicted the potential cessation of local mpox transmission in LAC in the absence of external introductions. Further details on calibration, validation, and model inputs can be found in Liang et al (18).

In the present study, we varied α ranging from 0.7 to 2.2 to reproduce observed case trends beginning in March 2023. We also used α to simulate the effects of public health interventions by lowering it during periods of interest to reflect reduced sexual activity and transmission risk.

#### Simulating Introductions and Integrating Phylodynamic Estimates

While the model suggests the potential cessation of local mpox transmission in LAC by March 2023, the empirical sporadic cases and minor surges observed afterward highlight the need to account for external viral introductions. To explore this, we first tested hypothetical scenarios in which 5, 10, or 15 symptomatic cases were introduced into the model each week. These cases were randomly drawn in proportion to the simulation demographic and were allowed to infect susceptibles in the model.

Simulation results showed that higher levels of constant weekly importations led to more local transmissions. However, these simplified scenarios did not reproduce the observed temporal patterns in mpox case counts, nor did they explain the persistence of low-level transmission (Fig. 1D). To better align the model with real-world dynamics, we integrated time-varying viral importation estimates from our phylodynamic analysis. By incorporating both the number and timing of imported cases more precisely, we improved the model’s ability to simulate short-term surges and temporal patterns in mpox case counts. Linking these external seeding events with behavioral changes modeled via the Infectivity Scalar (α), the combined framework helped explain how ongoing transmission can arise even after local transmission declined. As in the hypothetical scenarios, these phylodynamics-informed imported cases were treated as already diagnosed upon introduction, ensuring they were not counted as newly diagnosed within LAC.

#### Running the Model and Analyzing Output

Due to the stochastic nature of our model, we ran twenty independent iterations to capture inherent variability in the outcomes. To estimate uncertainty intervals for key metrics, such as incident case counts, we employed a bootstrap approach with 500 samples. Each sample consisted of a resample (with replacement) from the ten iterations. We calculated weekly averages for each sample, forming the data into a 500 × 85 matrix, where each row represents a bootstrap sample, and each column corresponds to a week. From this matrix, we computed the mean, lower bound (2.5th percentile), and upper bound (97.5th percentile) of these averages. This methodology captures the model’s stochastic variability and associated uncertainty in predicted outcomes. While we did not conduct a full probabilistic sensitivity analysis, we assessed the impact of parameter uncertainty through sensitivity tests on two alternative assumptions for vaccine efficacy, in which efficacy drops to 25% and 75%, compared to the base case 50% (see Fig. S14).

*Rt* was estimated by multiplying the average number of secondary infections per infected individual by the duration of infectiousness.

All simulations were programmed in MATLAB and executed on the high-performance computing facilities at the Center for Advanced Research Computing (CARC) (58). Using a parallel computing setup, we ran 20 iterations simultaneously, with each batch for a given scenario completing in approximately 13 minutes on a computing node with at least 20 available CPU cores. This approach allowed us to efficiently scale the number of iterations while maintaining a practical runtime for model calibration and scenario analysis.

### Phylodynamic Analysis

#### Genomic data and maximum likelihood tree generation

All available MPXV sequences were downloaded from GenBank on 01-20-2024. Sequences with ambiguous date of collection in the month column, with a sample collection earlier than January 2022, and flagged as being low quality by Nextclade https://docs.nextstrain.org/projects/nextclade/en/stable/user/algorithm/07-quality-control.html) (59) were excluded. Given that mpox transmission in the United States is driven by clade IIb viruses, sequences from other clades were also excluded, resulting in 7859 genome sequences included in our analysis.

A temporally-resolved phylogeny was created using a modified version of the Nextstrain (23) mpox workflow (https://github.com/nextstrain/mpox), which aligns sequences against the MK783032 (collection date: Nov. 2017) reference using nextalign (59), infers a maximum-likelihood phylogeny using IQ-TREE (60) with a GTR nucleotide substitution model, and estimates molecular clock branch lengths using TreeTime (61). The resulting phylogeny specific to this dataset can be found at https://nextstrain.org/groups/blab/mpox/allcladeIIseqs

#### Geographic scales

Due to the low number of sequences from various countries, we analyzed mpox spread with a focus on large metropolitan US cities and areas that have the highest level of mpox sequencing effort. Our focus areas were: Los Angeles County, California; Washington State; Cook County, Illinois; New York City, New York; California without Los Angeles County; North America excluding the areas previously mentioned; and Global regions outside of North America.

Given that Los Angeles County Department of Public Health (LA DPH) sequences the mpox cases for LAC, we assume that any genome labelled as being sequenced by LA DPH was sampled in LAC, while those sampled by the California Department of Health (CDPH) were sampled in locations within California but outside of LAC. From these 719 genomes, the dataset was filtered down to 497 by LA DPH to remove duplicated sequences from the same individual and samples that were collected outside of LA DPH. Despite this, there is always a small chance that CDPH might have received and sequenced a LAC case, but we expect this to be small and should result in a conservative bias as misclassification of an LAC sequence as non-LAC would result in smaller clusters and less intense transmission dynamics.

Phylogeographic reconstruction of mpox spread was conducted using the same Nextstrain workflow via ancestral trait reconstruction (62) of the aforementioned focus areas. This was done using the “augur traits” function (63).

#### Clustering

To identify local outbreak groups in Los Angeles County, we clustered all LAC sequences based on inferred internal node location. Following Müller et al (64) and Paredes et al (65), we used a parsimony-based approach to reconstruct the locations of internal nodes. Briefly, using the Fitch parsimony algorithm, we inferred internal node locations by considering only two sequence locations: LAC and then anywhere else. We then identified local outbreak clusters by selecting groups of sequences in which all their ancestral nodes were inferred to be from LAC, up until there was a change in location.

We then plotted the mean cluster size and the number of local clusters per month by using the month of collection for the first collected sequence of each identified outbreak cluster over time.

#### Estimating population dynamics jointly from multiple local outbreak clusters

To analyze the local transmission dynamics of mpox in LAC from 2022-2024, we used a multi-tree coalescent model to jointly model mpox dynamics from the inferred outbreak clusters, originally described in Müller et al (64). Briefly, we assumed that each identified cluster was the result of a single introduction into LAC and that the sequences that make up each cluster were the result of local transmission. Doing so allowed us to model mpox transmission as a structured coalescent process where the migration history is conditional on the clustering done *a priori.* The model allows mpox lineages to coalesce within LAC but can also originate from outside the sampled area. The migration history of the coalescent process is conditioned on the identified transmission clusters whereby we assume that the introduction event into LAC occurred prior to the most recent common ancestor of the sequences in each cluster. This time of introduction is explored via an MCMC run. We used a skyline approach to estimate both the effective population size (*Ne)* and rates of introduction throughout time using predefined change points (every 7 days), assuming exponential growth or decline between each change point. We ran two independent chains, and employed a strict molecular clock with a uniform distribution from 0 to 1 and a fixed value of 6 × 10^−5^ (6,23) and an HKY+Γ nucleotide substitution model with an estimated κ. We also repeat the analysis to test the sensitivity of our results with the following specifications: with a GTR+Γ substitution model with the same fixed clock rate and estimated frequencies and transitions; and with an eight-category discrete Γ prior instead of four (66).

Similar to Müller et al (64), we apply an exponential coalescent model with time-varying growth rates by accounting for correlations between adjacent *Nes* via the skyride approach, which assumes the log of adjacent *Ne* are normally distributed with a mean of 0 and an estimated variance. We also assumed the differences in growth rates were normally distributed with a mean of 0 and estimated variance. This formulation was validated in Supplementary Figure 3 of Müller et al (64).

#### Deriving and Implementing Case-Based Ne Prior

Additionally, we also conduct a separate analysis by allowing the Ne to be informed by the total number of diagnosed mpox cases in each month. In a standard formulation of the coalescent model of infectious diseases parameterized by Susceptible-Infected-Recovered (SIR) dynamics (67),

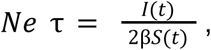

where τ refers to the generation time, *I*(*t*) and *S*(*t*) to the time varying prevalence and number of susceptibles in the population, respectively, and β to the transmission rate. We represent the scaler 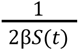 via θ(*t*) so that,

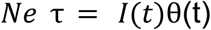

If we assume that the number of diagnosed mpox cases can approximate the prevalence *I(t)*, then we can rewrite the above equation as

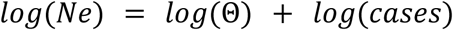

To account for time-varying observation noise and variability in the above assumptions, we can add an error term so that,

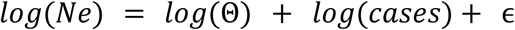

By rearranging the terms we get

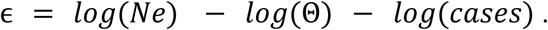

We then account for correlations between adjacent errors by assuming that the difference in errors is normally distributed with a mean of 0 and an estimated variance.

We implemented and ran these models as an extension to BEAST2 software version 2.6.2 (68) and can be found on https://github.com/miparedes/mab. We performed effective population size and migration rate inference using an adaptive multivariate Gaussian operator (69) and ran the analyses using an adaptive Metropolis-coupled MCMC (70) using two chains with a length of 2.5 × 10^8^. We repeat our analysis without the rolling mean smoothing as well as without any cases to test the sensitivity of our results.

### Posterior processing of Phylodynamic Analyses

Parameter traces were visually evaluated for convergence using Tracer (71), tree distributions were visually inspected using IcyTree (72), and 20% burn-in was applied for all phylodynamic analyses. All tree plotting was performed with baltic (https://github.com/evogytis/baltic) and data plotting was done using Altair (73), matplotlib (74) and seaborn (75).

Following Bedford et al. (76), persistence time was measured by calculating the average number of days for a lineage to leave LAC, walking backwards up the phylogeny from the tip up until the node location was outside of LAC. We also cycled through the posterior set of trees to find the median time of importation into LAC for each identified local outbreak cluster

#### Estimating number of imporations into LAC for Microsimulation Model

The absolute number of viral importation events into LAC was estimated by calculating the number of transitions walking from tips to root in the posterior set of trees and calculating the median as well as the 50% and 95% highest posterior density estimates (HPD).

#### Estimating percentage of new cases due to introductions

We estimated the percentage of new cases due to introductions for each global region by adapting the methods previously described (6). Briefly, the percentage of cases due to introductions π at time *t* can be calculated by dividing the number of introductions at time *t* by the total number of new cases at time *t.* We first represented the total number of new cases in a region as the sum of the number of introductions and the number of new local infections due to local transmission, resulting in the following equation:

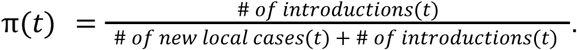

We estimated the number of new local cases at time *t* by assuming the local epidemic in each global region follows a simple transmission model, in which we derived the number of new cases at time *t* as the product of the transmission rate β (new infections per day per infected individual) multiplied by the number of people already infected in that region *I*. For the number of introductions, we similarly assumed that the number of introductions equals the product of the rate of introduction (introductions per day per infectious individual, which we refer to as migration rate *m*) and the number of people already infected in that region *I*. We use the number of infected individuals in the destination region rather than the origin region for calculating the number of introductions since the approximate structured coalescent approach models epidemic processes as backwards-in-time, resulting in the equation containing only information about the number of infected individuals in the destination region (more information on backwards migration rates below). We then rewrote the above equation as

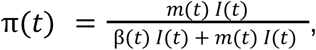

where *I*(*t*) denotes the number of infected people in that region at time *t*. Given the presence of *I*(*t*) in every element, we factored out *I*(*t*) to arrive at

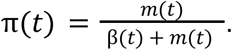

For each region, we considered introductions at time *t* to be the sum of the introductions coming into LA Country from outside the region. We define the percentage of new cases due to introductions π at time *t* for LAC as

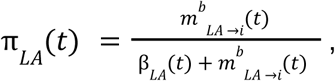

where *m^b^_LA →i_* denotes the backwards migration rate per lineage per day into LAC from outside and is estimated directly via our multi-tree coalescent model.

In a SEIR transmission modeling framework (employed due to the incubation period of MPXV), the transmission rate β is a function of the infectious period γ, the incubation period σ, and the exponential growth rate *r* (as adapted from Example 4 in Ma 2020 (77)):

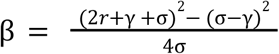

To compute the growth rate in region *y* for use in calculating the percentage of cases due to introductions only, we assumed that differences in effective population size between adjacent time intervals can approximate the growth rate *r* and thus 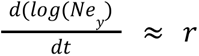. In addition, we assumed that 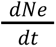 is independent from the rate of introduction. We calculated the growth rate of the effective population size 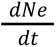 as

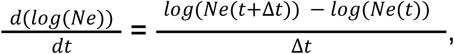

where *Ne*(*t*) denotes the effective population size of a region at time *t*. We ran our analysis using weekly time intervals but averaged over three week intervals ( Δ*t* = 3) for the growth rate in order to reduce noise and account for the long generation time for mpox.

We calculated the transmission rate β at time *t* in LAC as

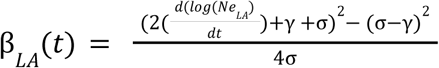

#### Incubation and infectious period estimates for use in phylodynamic analyses only

For the incubation period, we used a mean of 8 days based on prior literature (55,78). The infectiousness period for mpox has yet to be definitively characterized (79), as such we used the estimates of the infectious period (10.9 days) from Jeong et al (46) for our main analysis as they were defined via analysis of viral load and viral shedding in more than 90 mpox cases. To account for variability in this estimate, we also repeated our percentage of cases due to introductions and *Rt* analyses using a mean infectious period of 4.5 days and 21 days (Fig. S5). The mean infectious period of 4.5 days was estimated from the comparison of the generation time of 12.5 days (80) and the aforementioned incubation period through the formulation of the generation time in Wallinga and Lipsitch (81). This lower estimate of the infectious period is in concordance with the infectious period estimations from Zhang el at (82). The mean estimate of 21 days refers to the average time of resolution of symptoms (83) and has been previously used as a mainly-clinical proxy for infectiousness (84).

#### Estimating the effective reproductive number Rt from pathogen genomes

We calculated the effective reproductive number *Rt*, the time-varying average of secondary infections from a primary infected individuals, in LAC, assuming an exponentially distributed infectious and incubation period of mean respectively 1/γ and 1/σ, yielding 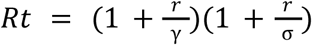 (81). Additionally, we sought to separate out the contributions of introductions versus local transmission to *Rt_t_*. To do so, we modified the *Rt* equation to include the percent of new cases from introductions as an estimate of local community spread so that 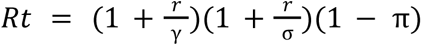, where π refers to the percentage of new cases due to introductions as described above.

Of note, our *Rt* calculations assume that the change in *Ne* over time is proportional to the change in the number of infected individuals over time.

To further validate our estimates of *Rt,* we fit the estimate cluster distributions taken from the sizes of the identified sequenced outbreak clusters to the formulation in Tran-Kiem & Bedford (25) which allows for the estimation of *R* and the dispersal parameter *k* and accounts for the probability of a case being detected and sequenced (similar to (27,85)). Given that we use all available sequences and not just identical sequences, we set the probability that a transmission event occurs before a substitution event *p* as 1. We also assume a range of case detection rates from 5% to 100% of all cases detected and then sequenced. We report the results assuming a 5% case detection rate as the most conservative estimate. Similarly, we also explored the probability to observe at least a cluster of size 16 (the largest size found in 2023-2024) among 64 total clusters as a function of the effective reproduction number *R* across a time period, transmission heterogeneity as estimated via the dispersion parameter *k*, and the fraction of infections sequenced. This estimation has been previously derived in other work (24,25). We explore this probability among R values ranging from 0.1 to 1.6 and k values from 0 to 10, assuming a probability of case detection of 5.5% which was estimated to be the average case sequencing rate throughout the 2022 mpox epidemic (6). While we expect the fraction of infections sequenced to be higher in LAC for 2023-2024 (Fig S1), we use 5.5% as a conservative estimate, as increasing the fraction sequenced is likely to make even lower R values more likely.

#### Estimating the relative importance of introductions compared to ongoing lineages

To estimate the relative success of introductions over two month time windows from March 1, 2023 through December 12, 2024 (the date of the last sequence in our sample), we adapted the methods in Lemey 2021 (86). Briefly, we analyzed the posterior set of trees from our unstructured coalescent analysis to estimate the posterior mean and 95% HPDs of three proportions: the proportion of unique introductions in the time period over the total number of unique persisting lineages and unique introductions; the proportion of unique introductions whose downstream transmission chains persisted at least until the end of that time period over the total number of persistent introductions and ongoing lineages; and the proportion of descendant lineages from these unique introduction events over the total number of descendants circulating after the end of the two month time period. We define a unique introduction as the inferred migration date of an identified outbreak cluster that falls between the start and end date of each two month time slice. We define an ongoing lineage as a lineage whose inferred date of introduction is before the start date of each time period and exists through the end date. Similarly we define descendants as the number of leaves (sequences) stemming either from the unique introductions or the ongoing lineages that exist at or after the end date of each time period.

#### Phylodynamic Simulations

To test the applicability of our multitree coalescent model both with the standard implementation as well as our cases-informed *Ne*, we simulated phylogenetic trees under an SEIR model with superspreading (64). We also assumed a constant force of introduction per unit time into the region. We assumed the number of newly infected individuals to be negatively binomially distributed such that the mean number of introductions at any point in time *t* was equal to *Rt* and the dispersion parameter *k* = 0.3 as previously estimated (6,26). To approximate real-life sampling dynamics, we parameterized the sampling rate based on the estimated time to present to healthcare in the UK in 2022 (87). We next simulated a structured phylogenetic tree from this approach and then simulated genetic sequences on top of this phylogenetic tree using Seq-Gen (88) assuming an HKY substitution model, a genome size of 197,000bps and a clock rate of 6 × 10^−5^, similar to our main analysis above. To understand the impact of undersampling, we also randomly subsampled 50% of the simulated sequences and ran all the simulations via our multi-tree coalescent models. We then compared the estimated *Ne, Rt,* and percentage of cases due to introductions with the same values calculated from the SEIR dynamics.

## Data and Code Availability

Nextstrain builds, BEAST2 XMLs, scripts, sequence information, and de-identified data for the phylogenetic and phylodynamic analyses can be found at https://github.com/blab/mpox-la/tree/main. All sequences are available on GenBank with accession numbers found in the supplementary information. The code for the microsimulation model developed to study mpox incidence and dynamics is available at https://github.com/citina/microsimulation-mpox, which includes all scripts, parameter files, and usage instructions necessary to replicate the study findings.

## Acknowledgements

We would like to thank Cécile Tran Kiem for helpful discussions surrounding estimating epidemiological parameters from cluster distributions.The authors would like to thank Peera Hemarajata and staff in the Molecular Epidemiology, Bioterrorism Response, and Sequencing units at Los Angeles County Public Health Laboratories for their technical assistance. We gratefully acknowledge all data contributors, i.e., the authors and their originating laboratories responsible for obtaining the specimens, and their submitting laboratories for generating the genetic sequence and metadata and sharing via GenBank. We have included a detailed acknowledgment table in https://github.com/blab/mpox-la.

## Author Contributions

Conceived and designed the study: MIP, CL, SCS, IWH, NFM, JO

Collected the data: JMG, NGM,

Conducted the analysis: MIP, CL

Advised on analysis: SCS, IWH, TB, NFM, JO

Drafted the manuscript: MIP, CL, IWH, JO

Reviewed and edited the manuscript: All authors

## Ethics Approval

All data used in this study is publicly available, suitably anonymized viral sequence or epidemiological data. As such it does not constitute human-subjects research.

## Supplementary Material

**Figure S1:**
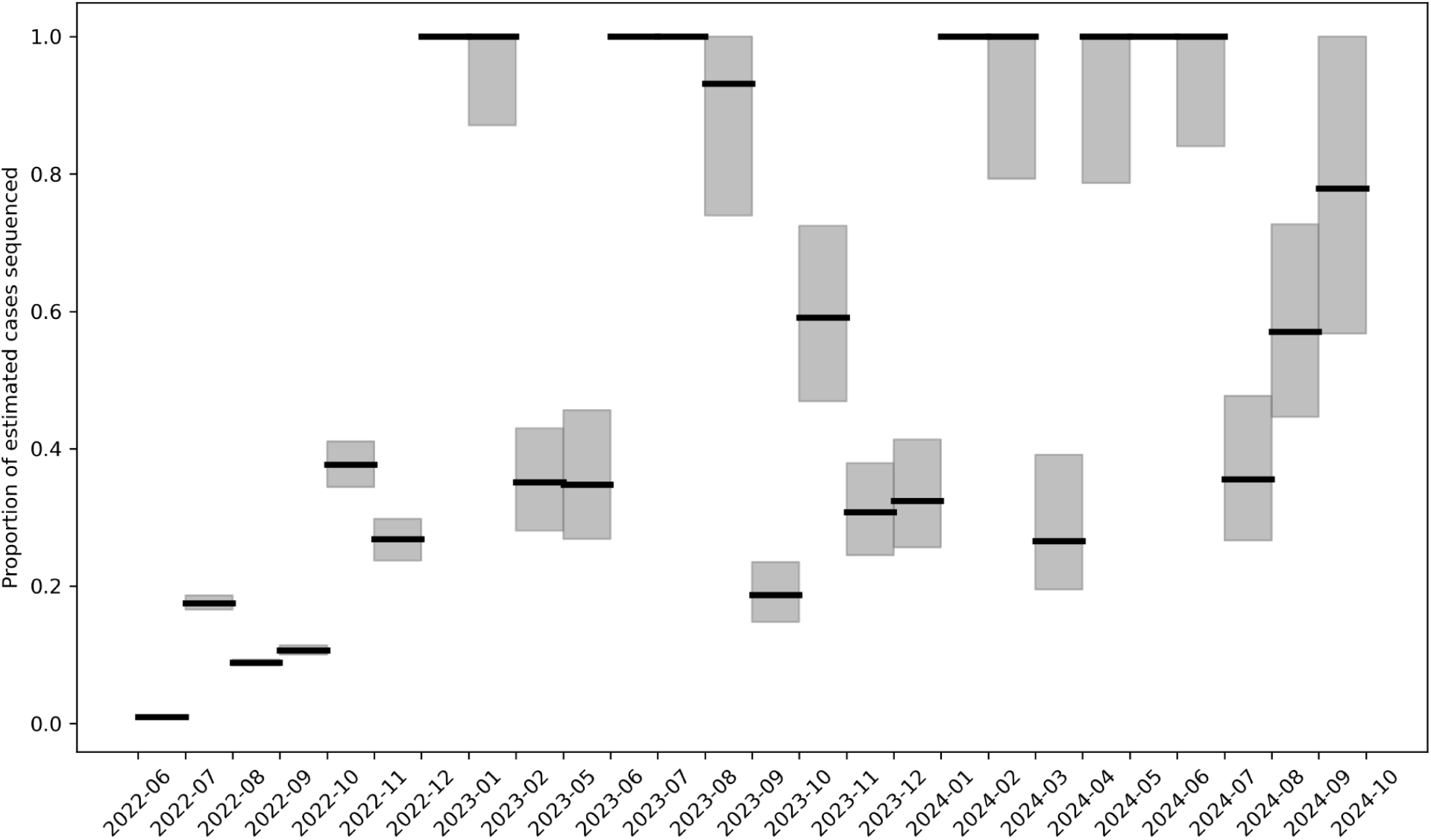
Estimated proportion of mpox cases sequenced in Los Angeles County. The proportion of cases sequenced was calculated by dividing the total number of mpox sequences from LAC found on GenBank by the monthly mpox incidence estimated from case counts using a renewal equation framework. The dark black horizontal line refers to the median estimates with the grey bars representing the 95% CI based on uncertainty in the incidence estimates. Months where the estimated proportion was greater than 100% (due to uncertainty incidence estimation due to low case counts or sample collection at dates different than diagnosis) were bounded at 100%.

**Figure S2:**
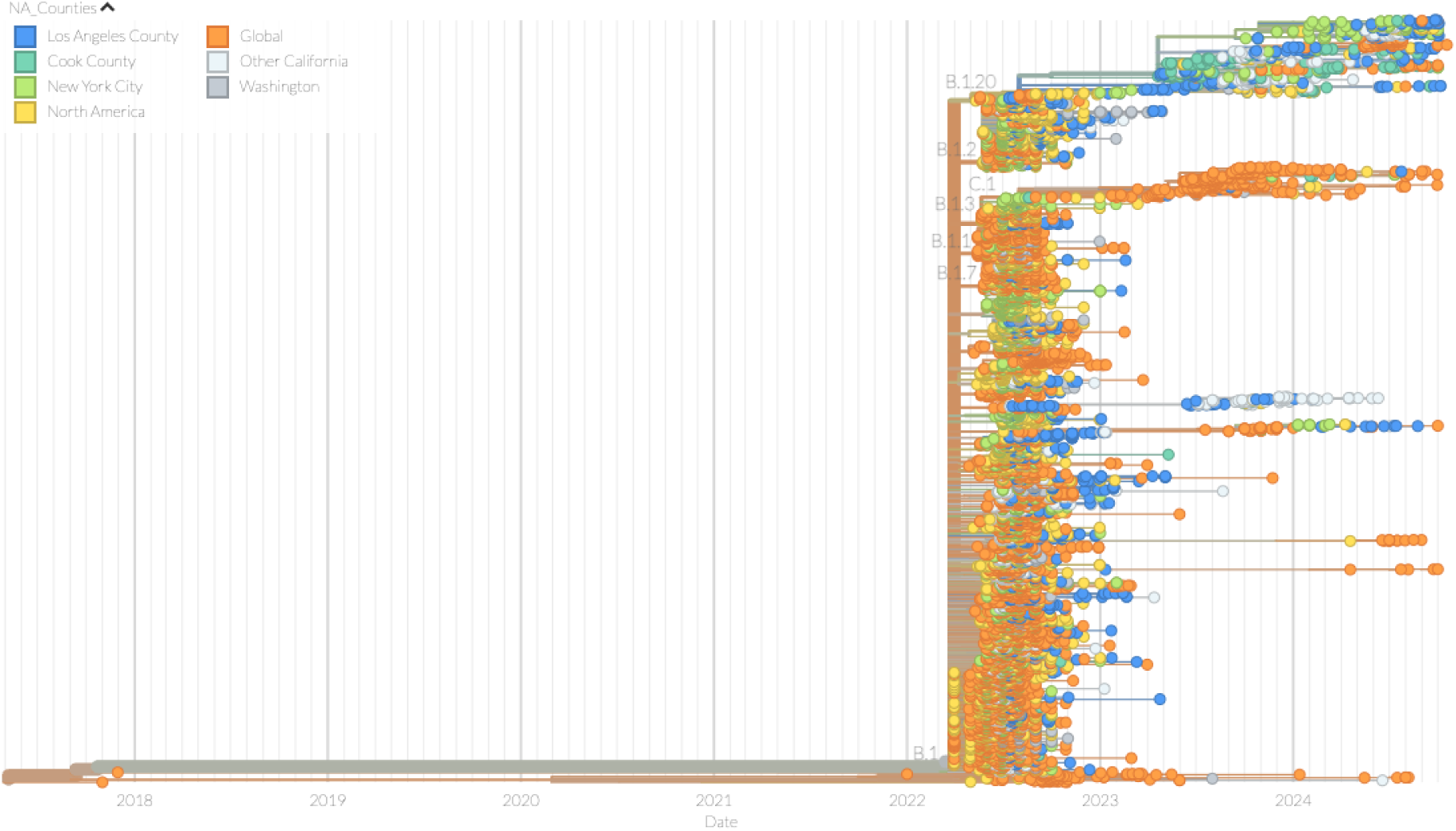
Time-resolved maximum likelihood phylogenies for mpox clade IIb sample collection dates between January 1 2022 and December 12, 2024. Tip color represents the focus areas in North America with high sequencing effort. Branches are colored based on inferred ancestry. The full tree can be explored interactively at https://nextstrain.org/groups/blab/mpox-la/allcladeIIseqs?c=focus_areas

**Figure S3:**
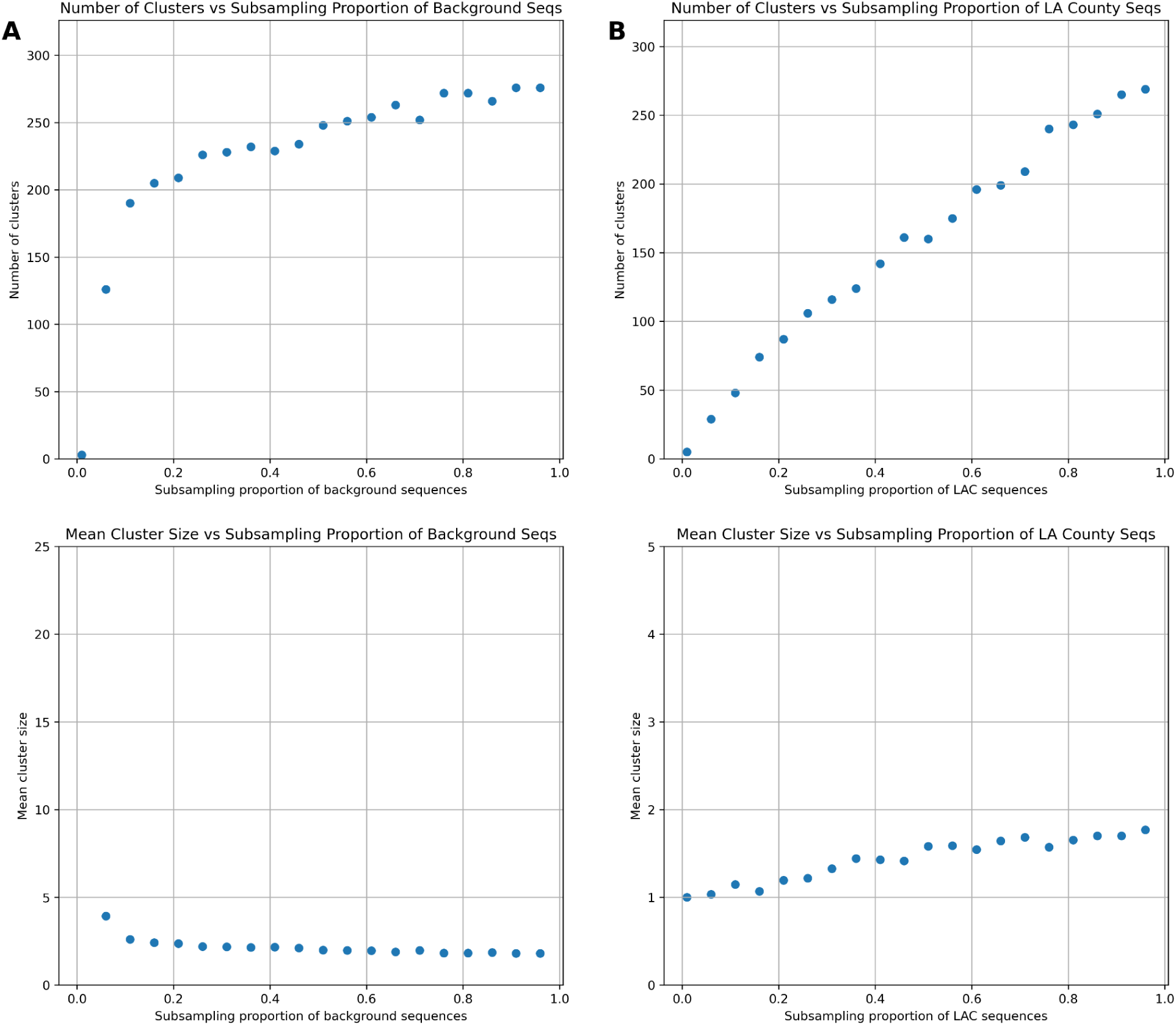
The impact of subsampling on the number and size of transmission clusters identified. We randomly subsampled different proportions of background sequences **(A)** and sequences from LA **(B)** and ran our clustering algorithm to show the impact of increasing the proportion sequences relative to the full dataset on the total number of clusters identified (Top row) and the mean size of those clusters (bottom row).

**Figure S4:**
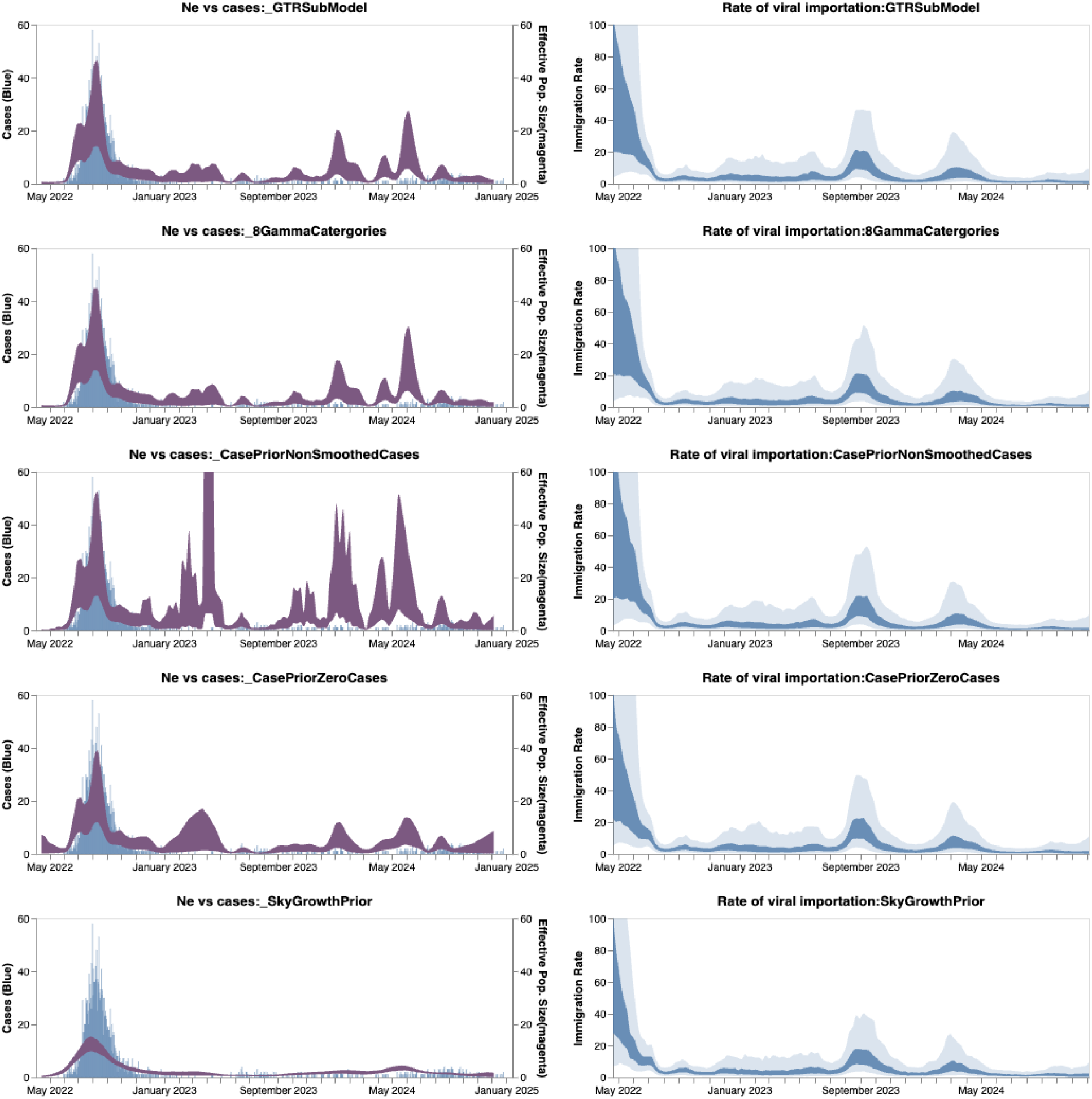
Sensitivity analysis of phylodynamic results: We tested the robustness of our phylodynamic results by repeating our main analyses under different model specifications. The left column shows the empirical case counts in blue and the estimated effective population size (Ne) in magenta (50% HPD). The right columns the inferred immigration rate of mpox into LAC, with the dark blue band representing the 50% HPD and the lighter blue representing the 95% HPD. The first row represents the same case-informed estimates as our main result but with a GTR substitution model instead of HKY. Second row represents 8 category discretization of the gamma distribution prior instead of 4 categories. The third row is our main model but without the cases being smoothed prior to being inputted into the model. The fourth row is a skyline prior represented by having zero case information in the case prior. The final row is a skygrowth prior with no case information instead of the skyline case-informed prior.

**Figure S5:**
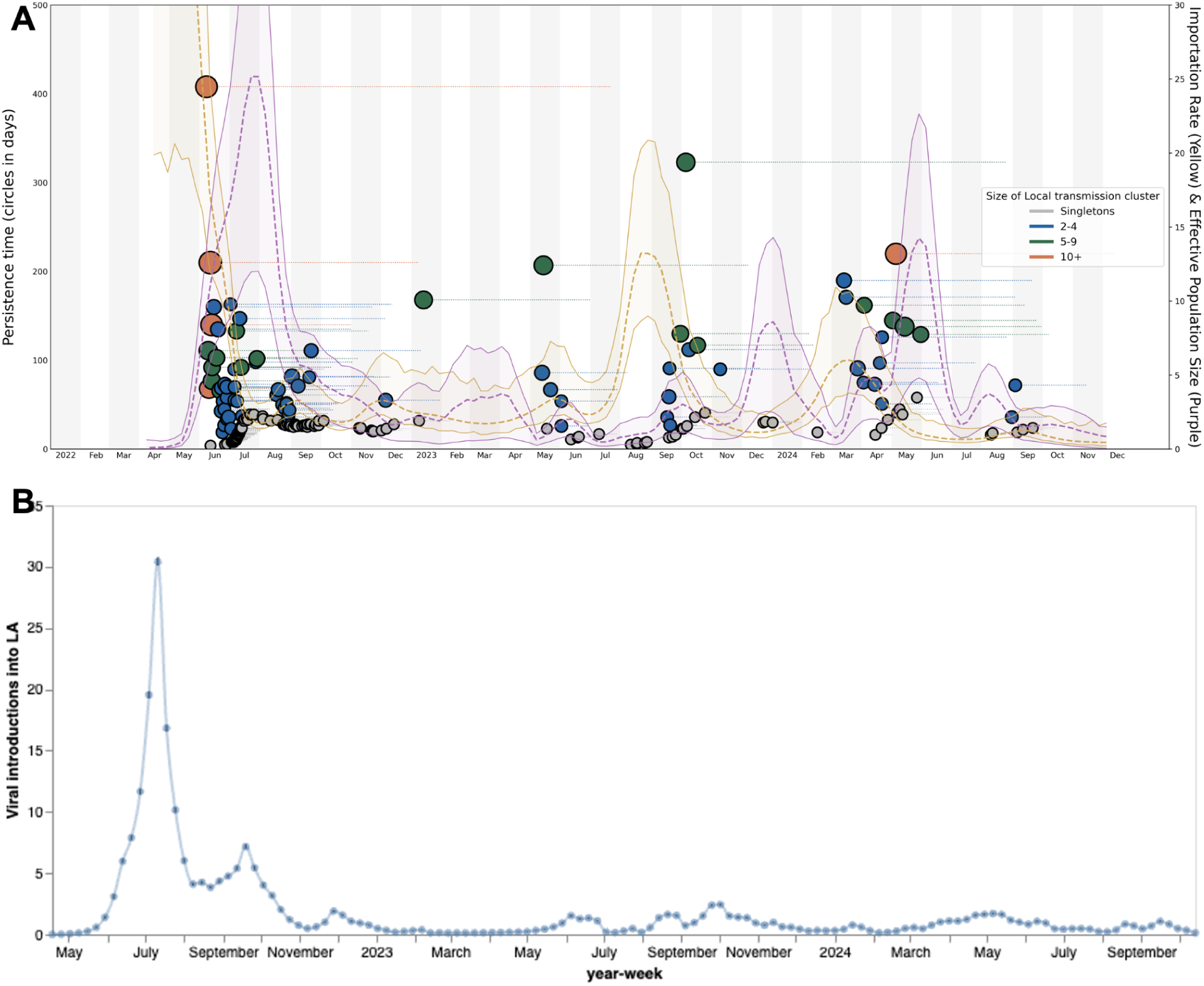
Mpox importation dynamics in LA County estimated via Bayesian Phylodynamics. Panel **A** shows the persistence time of each identified local outbreak cluster according to the date of its inferred introduction time. Each dot represents an inferred introduction into LA County, the radius of the dot is proportional to the size of the resulting transmission cluster. The yellow streamgraph is the rate of introduction (events/lineage/year) into LA county and the purple streamgraph represents the estimated effective population size, both inferred by our phylodynamic model. The dashed line represents the median and the bands represent the 95% HPD. Panel **B** shows the absolute number of viral introductions inferred via our phylodynamic model for each week that was calculated by analyzing the entire posterior set of phylogenetic trees after burn in. The error bars represent the 95% CI and these estimates were used to parameterize our microsimulation model.

**Figure S6:**
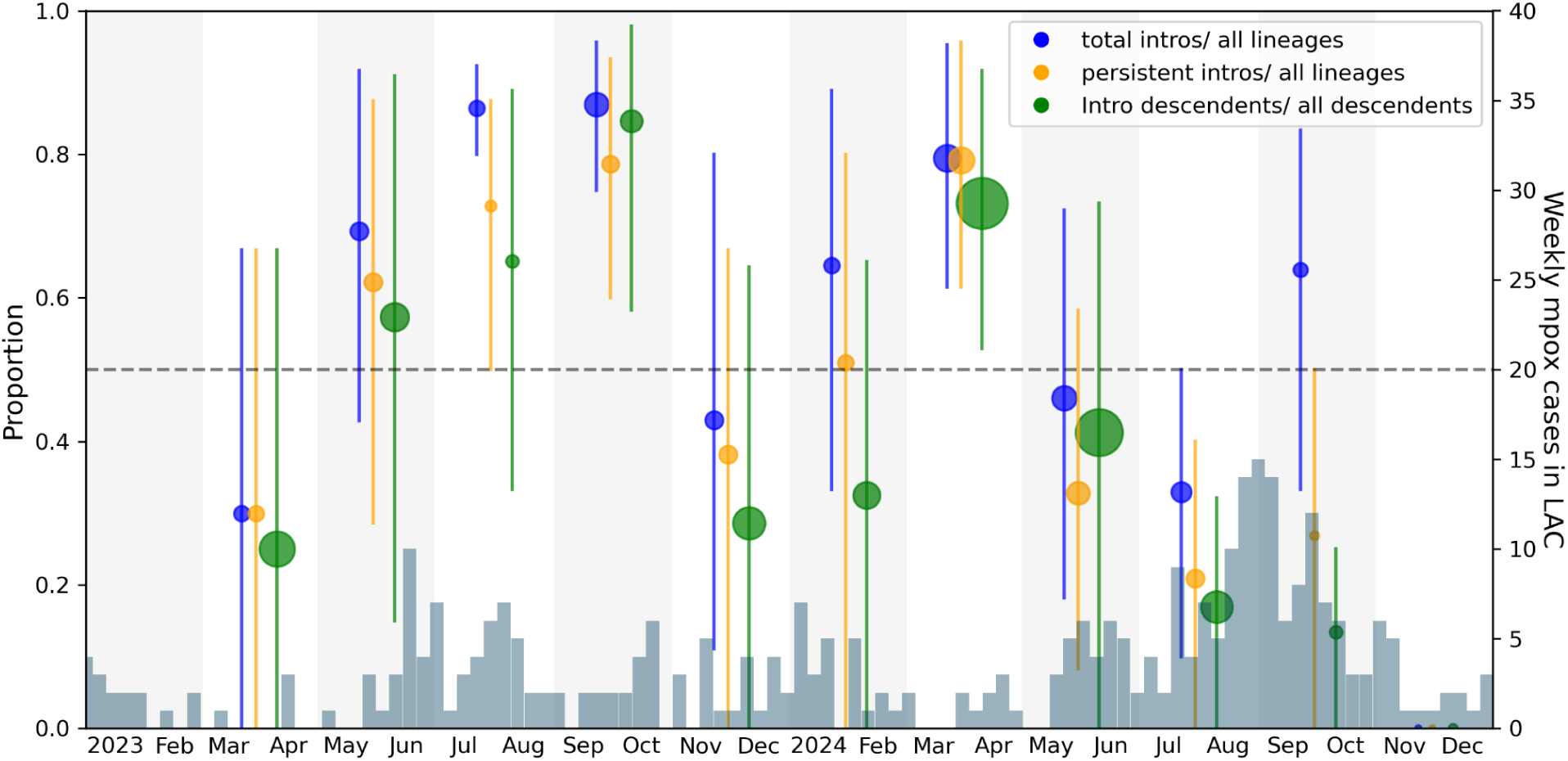
Posterior estimates of relative impact of introductions in LAC every two months from March 2023-December 2024. For every two months from March 1, 2023 through December 12, 2024 (denoted by alternating white and light gray vertical stripes), we report three posterior means and 95% HPD intervals for the relative impact of introductions in LAC estimated using the posterior set of phylogenetic trees from our unstructured coalescent model: (**Blue)** the proportion of unique introductions in the time period over the total number of unique persisting lineages and unique introductions; **(orange)** the proportion of unique introductions whose downstream transmission chains persisted at least until the end of that time period over the total number of persistent introductions and ongoing lineages; and **(green)** the proportion of descendant lineages from these unique introduction events over the total number of descendants circulating by the end of the two month time period. The points are scaled proportional to the total number of lineages or descendants in each time period that respectively contributed to the proportion. The gray bars represent the weekly cases in LAC. Given that our most recent sequence as on Dec 12th 2024, no viral introductions were found in Nov-Dec 2024 as more samples and time are needed to observe successful introductions.

**Figure S7:**
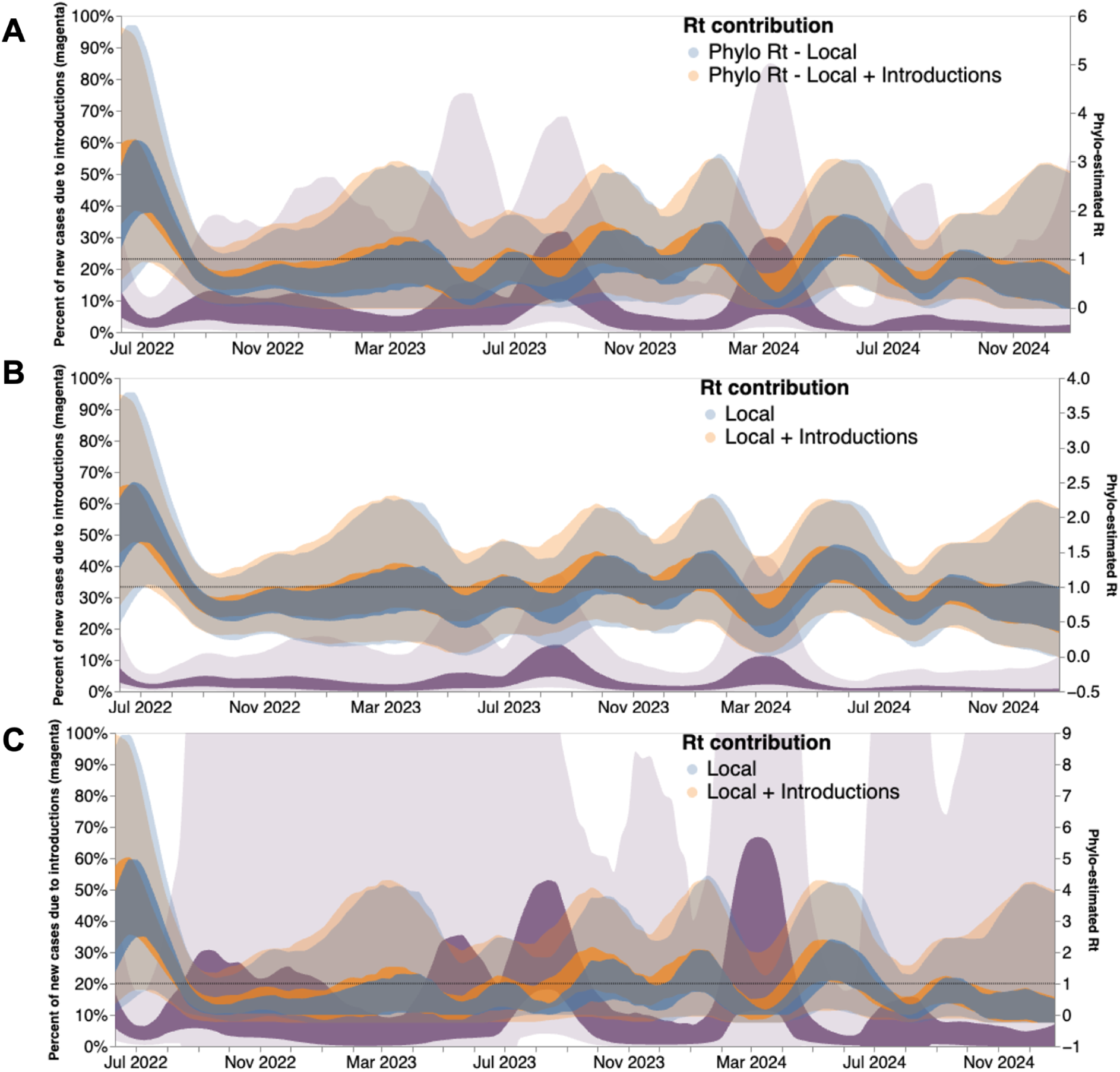
Percentage of new cases due to introductions and *Rt* with infectious period of (A) 10.9, (B) 4.5 and (C) 21 days. The inner area denotes the 50% HPD interval, and the outer area denotes the 95% HPD interval. The blue and orange bands lines represent estimates of *Rt* highlighting the contribution of local transmission only (blue) as well as that of viral introductions (orange). Dashed line highlights an *Rt* value of 1. *Rt* estimates were smoothed using a 14-day rolling average.

**Figure S8:**
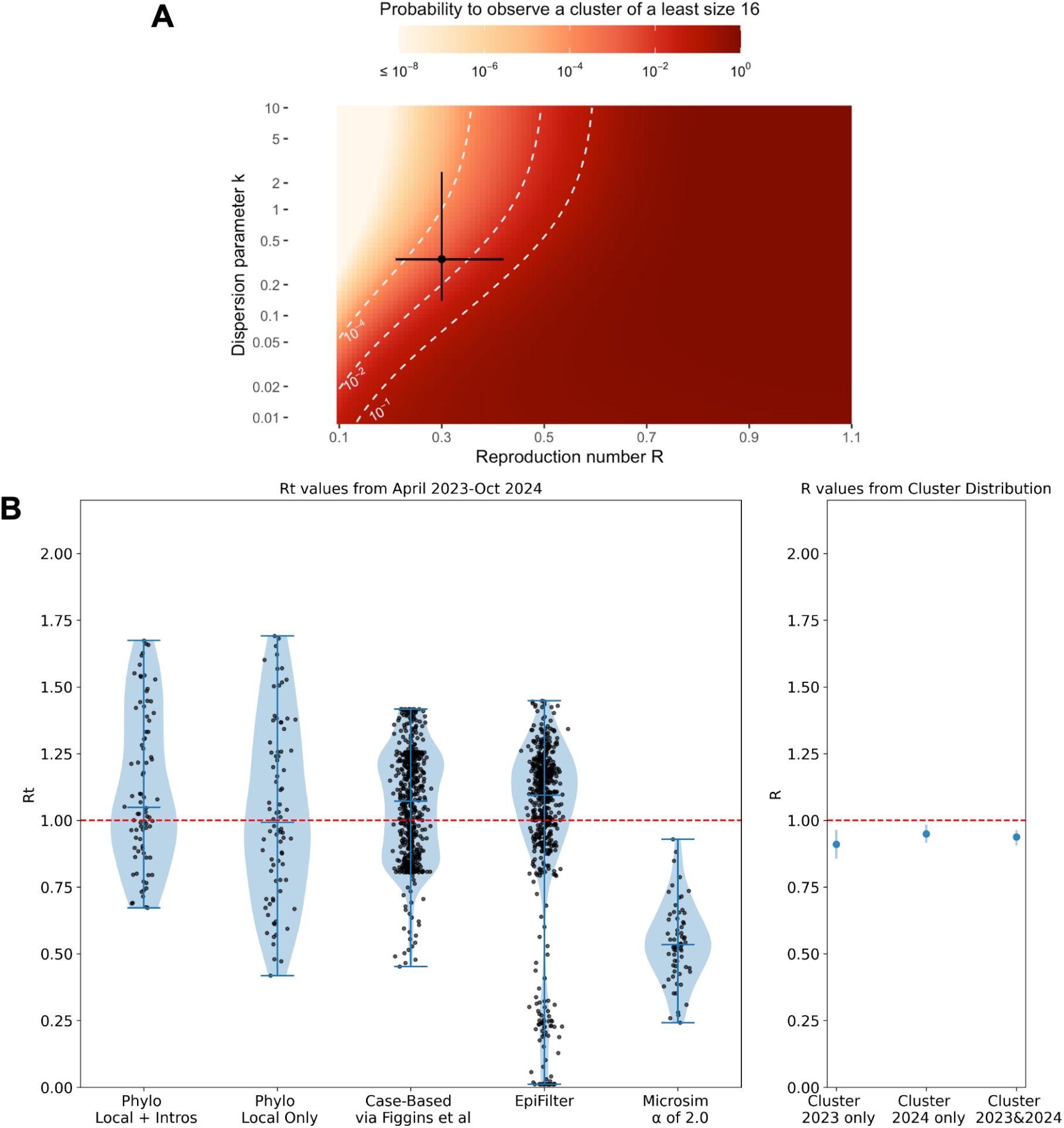
Reproductive number estimation for LAC from March 2023 through October 2024. **A.** Probability to observe a cluster of size 16 among 64 clusters as a function of the reproduction number R and the dispersion parameter k assuming 5.5% of infections are sequenced. The horizontal and vertical lines correspond to estimates obtained by Blumberg and Lloyd-Smith (27) from the analysis of epidemiological clusters during previous outbreaks. The dotted white lines correspond to contour lines for probabilities of 10−4, 10−2, and 10−1. **B.** The mean estimates of *Rt* (left) or R (right) for mpox showing the spread via a violin plot with the extremes and the median highlighted by the darker blue horizontal lines for diverse methodologies. The left panel plots the spread of weekly *Rt* estimations while the right panel shows the estimates of R with 95% CIs found from the distribution of cluster sizes for either 2023, 2024, or both years combined. The x axis of the left panel shows the methodology used and the dashed red line denotes an R or *Rt* of 1.

**Figure S9:**
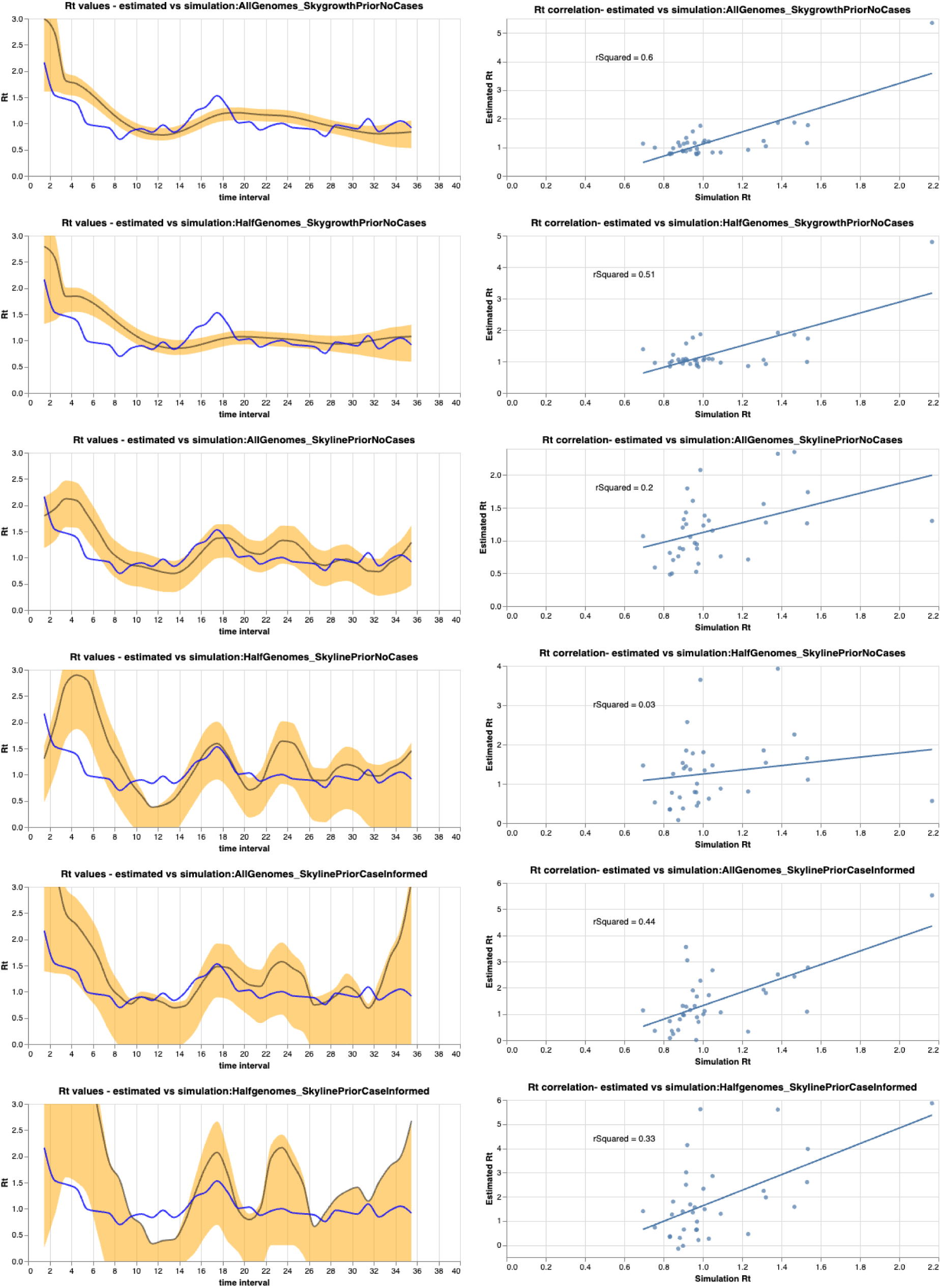
Estimation of *Rt* from simulations. We tested the ability of our multitree coalescent approach under different Ne priors to correctly estimate *Rt* from simulations. To do so, we simulated a local outbreak using a constant rate of introduction. We then sampled cases based on the estimated time to seek care during the 2022 mpox epidemic, subsampled, simulated genetic sequences, and then used the local transmission cluster to estimate Rt. The left column shows the true *Rt* in blue with the estimated *Rt* in yellow (showing the 95% HPD intervals) with the grey line representing the median estimate. The right column shows the correlation between simulated and the estimated *Rt* with a linear regression fit and R^2^ calculated. The first two rows represent a skygrowth prior on growth rate, the second two represent a skyline prior on the Ne without cases, and the bottom set of two represent the main analysis of a skyline prior on the Ne informed by mpox cases. For each set of priors, the top analysis represents 100% of the sampled genomes used while the bottom analysis represents only 50% of the genomes used. Estimates were smoothed using a 14 day rolling average.

**Figure S10:**
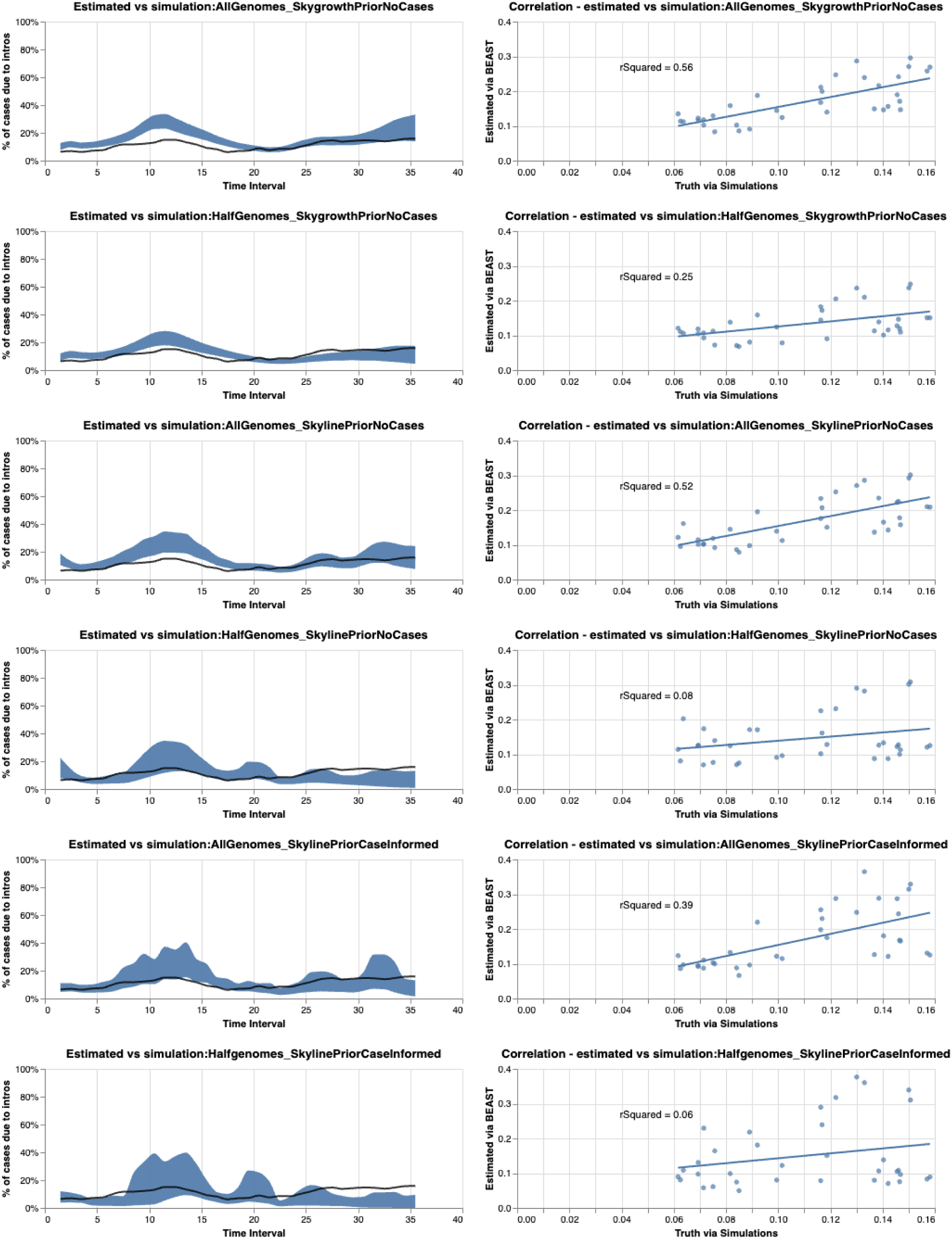
Estimation of percentage of cases due to introductions from simulations. We tested the ability of our multitree coalescent approach under different Ne priors to correctly estimate the percentage of cases due to introductions from simulations. To do so, we simulated a local outbreak using a constant rate of introduction. We then sampled cases based on the estimated time to seek care during the 2022 mpox epidemic, subsampled, simulated genetic sequences, and then used the local transmission cluster to estimate percentage The left column shows the true percentage in blue with the estimated percentage in yellow (showing the 95% HPD intervals) with the grey line representing the median estimate. The right column shows the correlation between simulated and the estimated percentage with a linear regression fit and R^2^ calculated. The first two rows represent a skygrowth prior on growth rate, the second two represent a skyline prior on the Ne without cases, and the bottom set of two represent the main analysis of a skyline prior on the Ne informed by mpox cases. For each set of priors, the top analysis represents 100% of the sampled genomes used while the bottom analysis represents only 50% of the genomes used. Estimates were smoothed using a 14 day rolling average.

**Figure S11:**
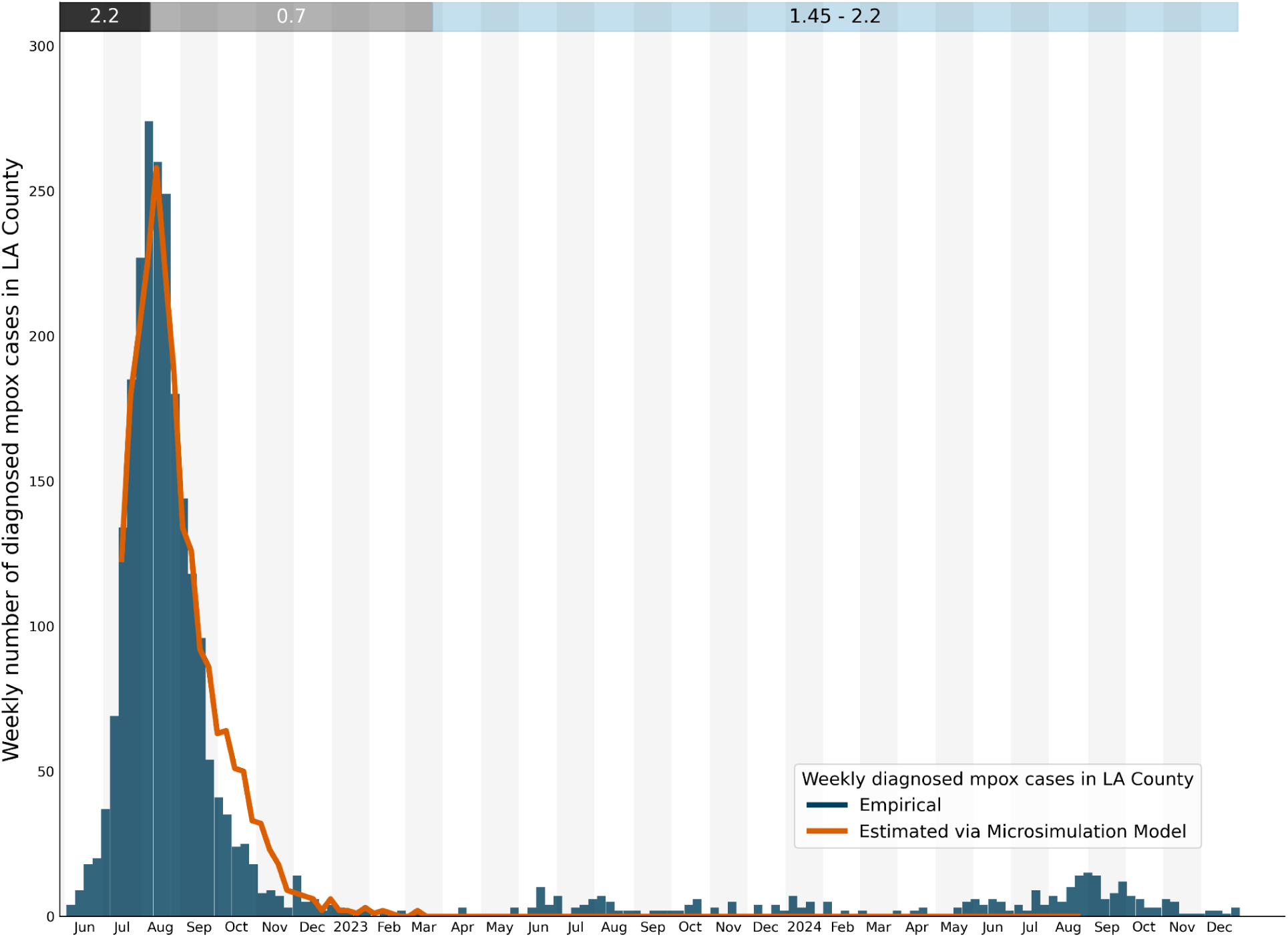
Infectivity Scalar (α) during 2022 mpox outbreak in Los Angeles County. Main figure shows the weekly number of diagnosed mpox cases in LAC from June 2022 through December 2024 (blue) with the number of diagnosed mpox cases simulated via our microsimulation model without viral importations overlaid in orange. The bars in the top of the figure are a visual representation of the periods of time for which α was calibrated. The grey bars represent the initial model calibration for the epidemic period and the blue bar shows the period of interest for this study.

**Figure S12:**
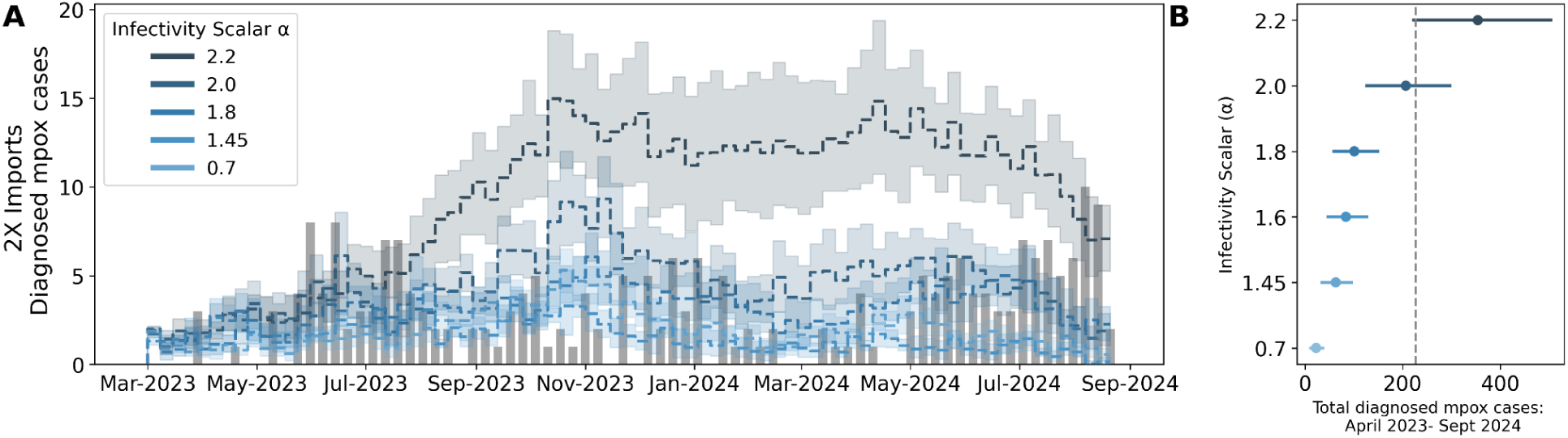
Infectivity Scalar α with twice as many phylodynamics-informed viral introductions. To test the impact of underestimating the number of viral introductions into LAC, we doubled the number of introductions, reran our microsimulation model, and explored the Infectivity Scalar α that best explains the empirical weekly number of diagnosed mpox cases (gray bars). Line graphs represent the mean weekly number of mpox diagnoses simulated using increasing α. Each weekly estimate represents the average of 10 independent iterations of our model. The dashed line in **B** represents the total empirical number of diagnosed mpox cases from April 2023 through September 2024.

**Figure S13:**
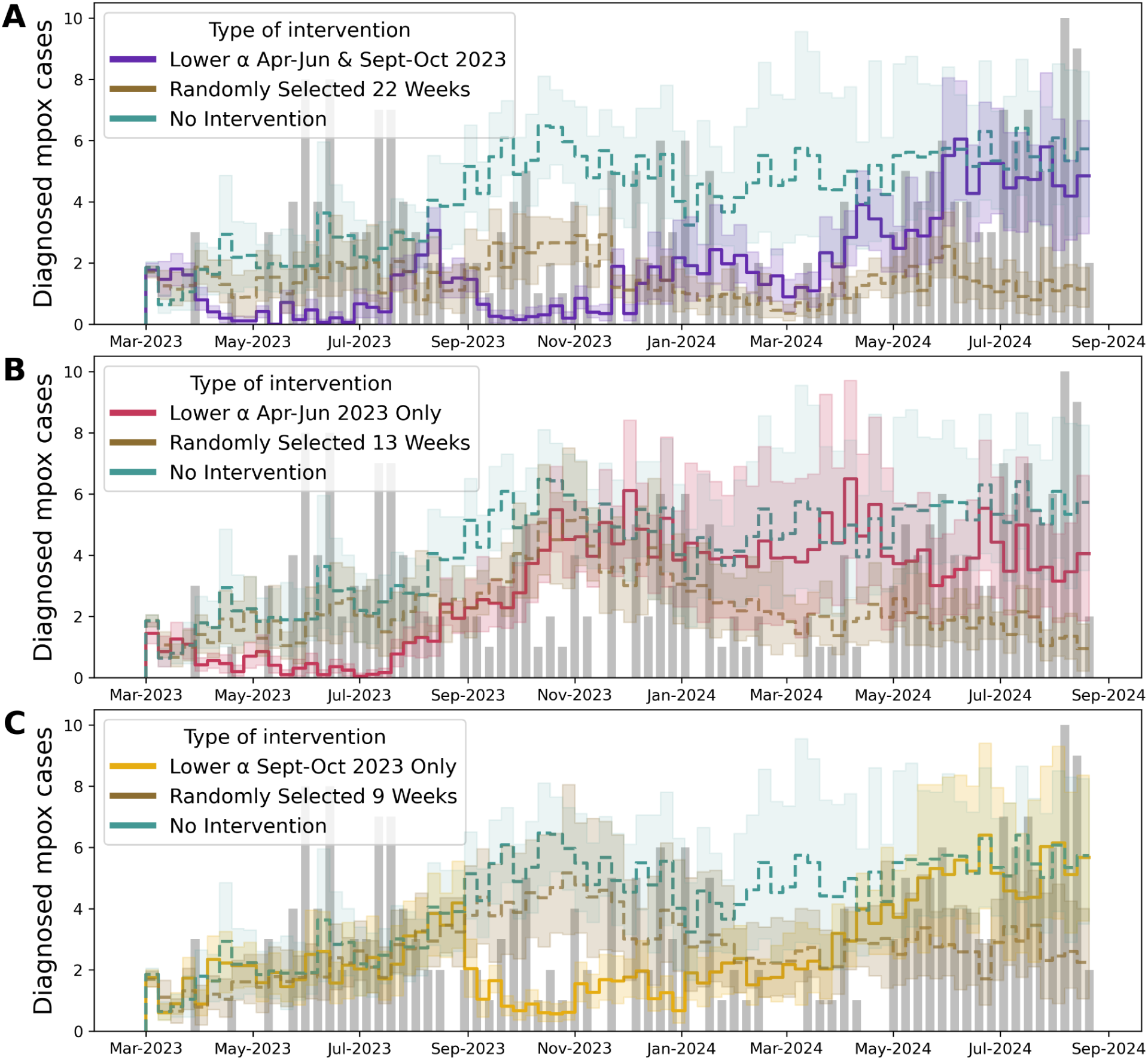
Factors maintaining mpox prevalence and modeling counterfactual public health interventions with statistical tests. **(A-C)** Given the non-constant pattern of viral introductions seen in the phylodynamic analysis, we tested different counterfactual scenarios of public health interventions during specific time periods represented by lowering the α to 0.7 while keeping the α at 2.2 during the remaining time. The bold yellow, red, and purple solid lines represent the simulated weekly number of diagnosed mpox cases under phylodynamic-informed interventions. To test for non-specific effects, we also reran our microsimulation model by randomly selecting the same number of weeks as our phylodynamics-informed interventions to lower the α to 0.7 (brown dashed lines) as well as a simulation without any interventions. (green dashed lines) In all plots, the grey bars represent the empirical number of mpox diagnoses in LAC.

**Figure S14:**
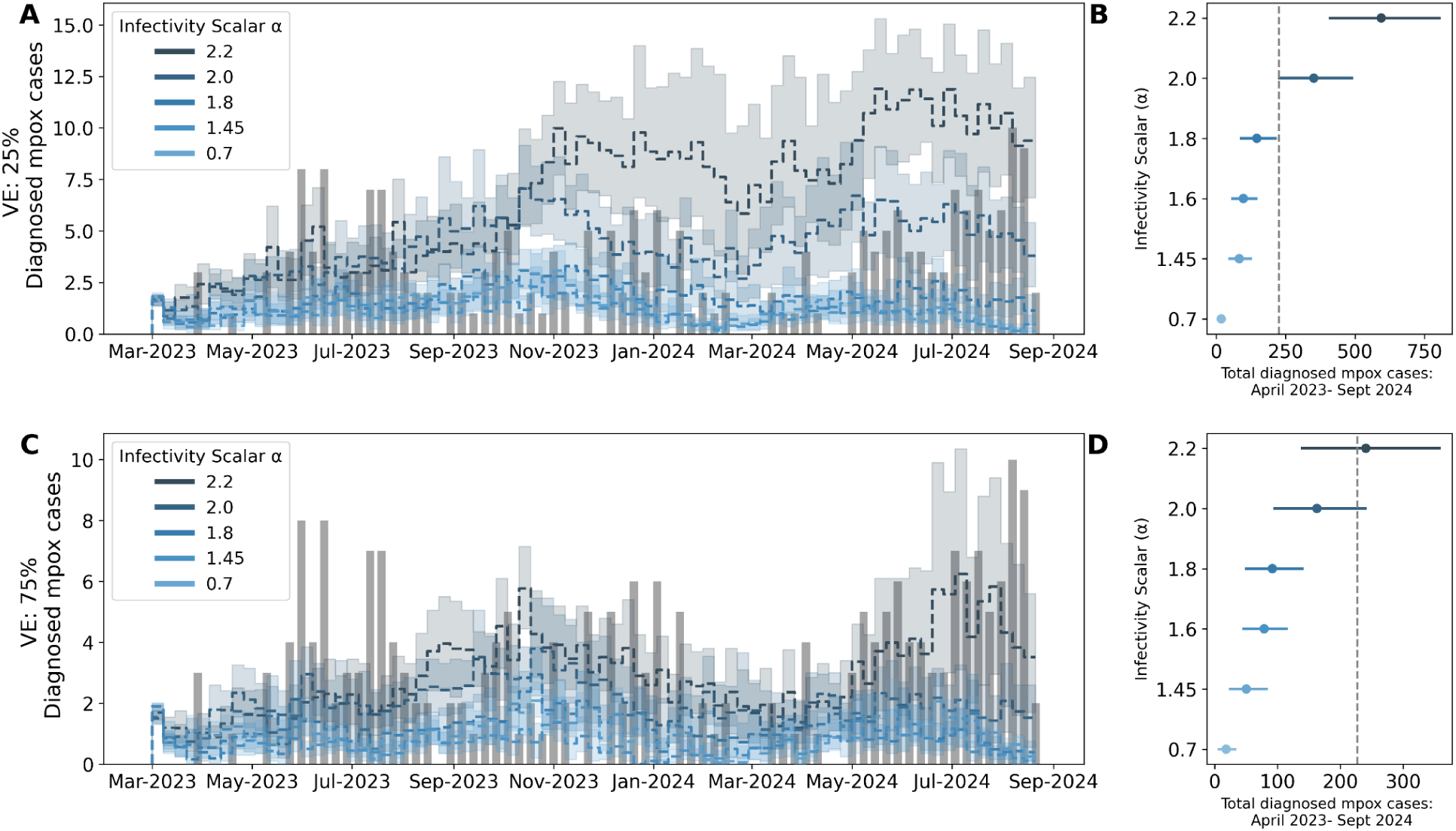
Sensitivity analysis of the impact of vaccine immune waning on α calibration. Here we repeat the analysis in Main Figure 5A-B, with the vaccine effectiveness against infection declining to 25% (A-B) and 75% (C-D) after one year. Line graphs **(A)** represent the mean weekly number of mpox diagnoses simulated using increasing α. The dashed line in **B** represents the total empirical number of diagnosed mpox cases from April 2023 through September 2024.

**Figure S15:**
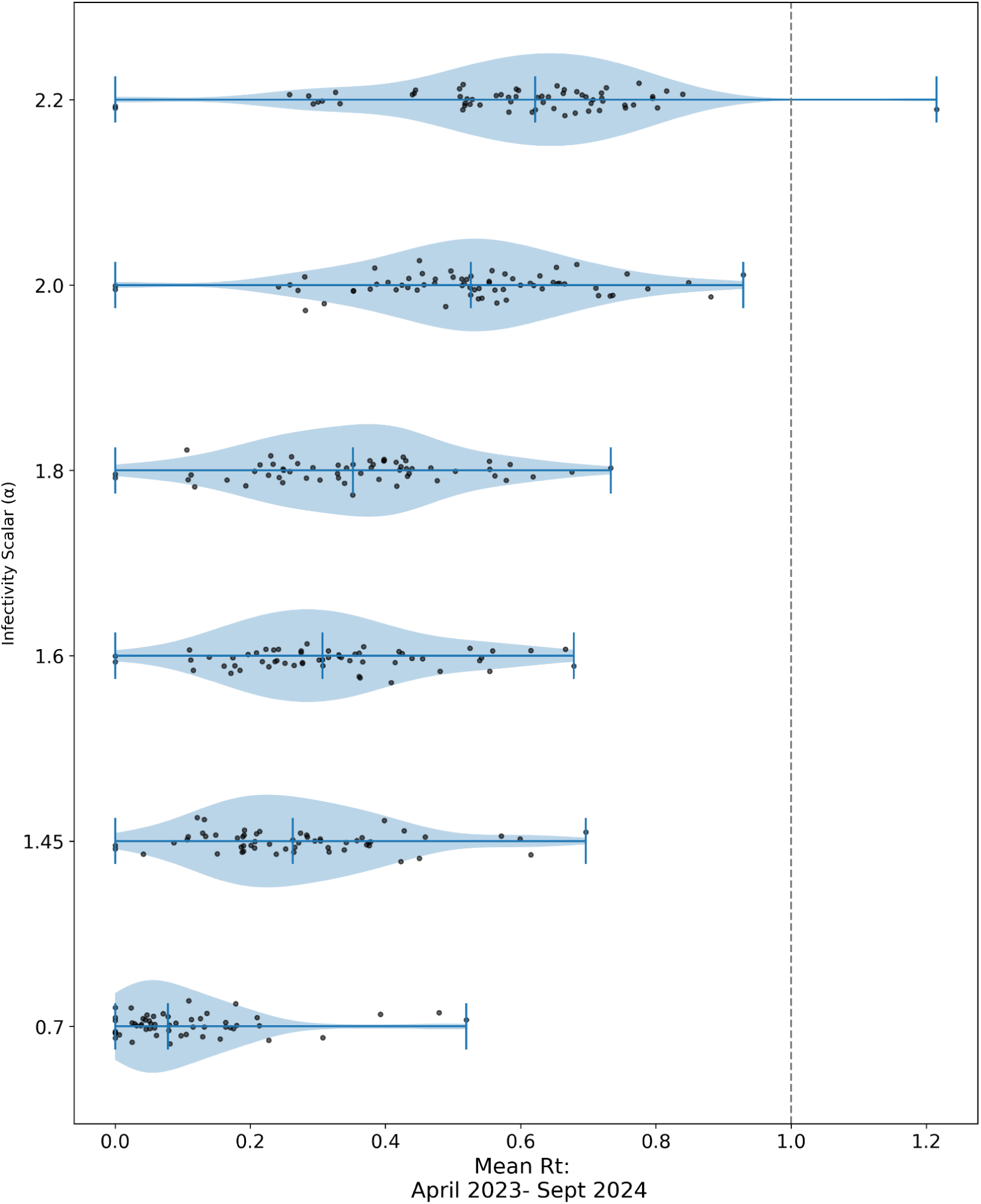
Mean *Rt* estimates for varying values of the Infectivity scalar via microsimulation for LAC from March 2023 through October 2024. The mean estimates of *Rt* for mpox for increasing values of α showing the spread via a violin plot with the extremes and the median highlighted by the darker blue horizontal lines. The dashed black line denotes an *Rt* of 1. *Rt* was estimated by tracking the average weekly number of secondary infections per infected individual multiplied by the time that individuals remain infectious.

**Figure S16.**
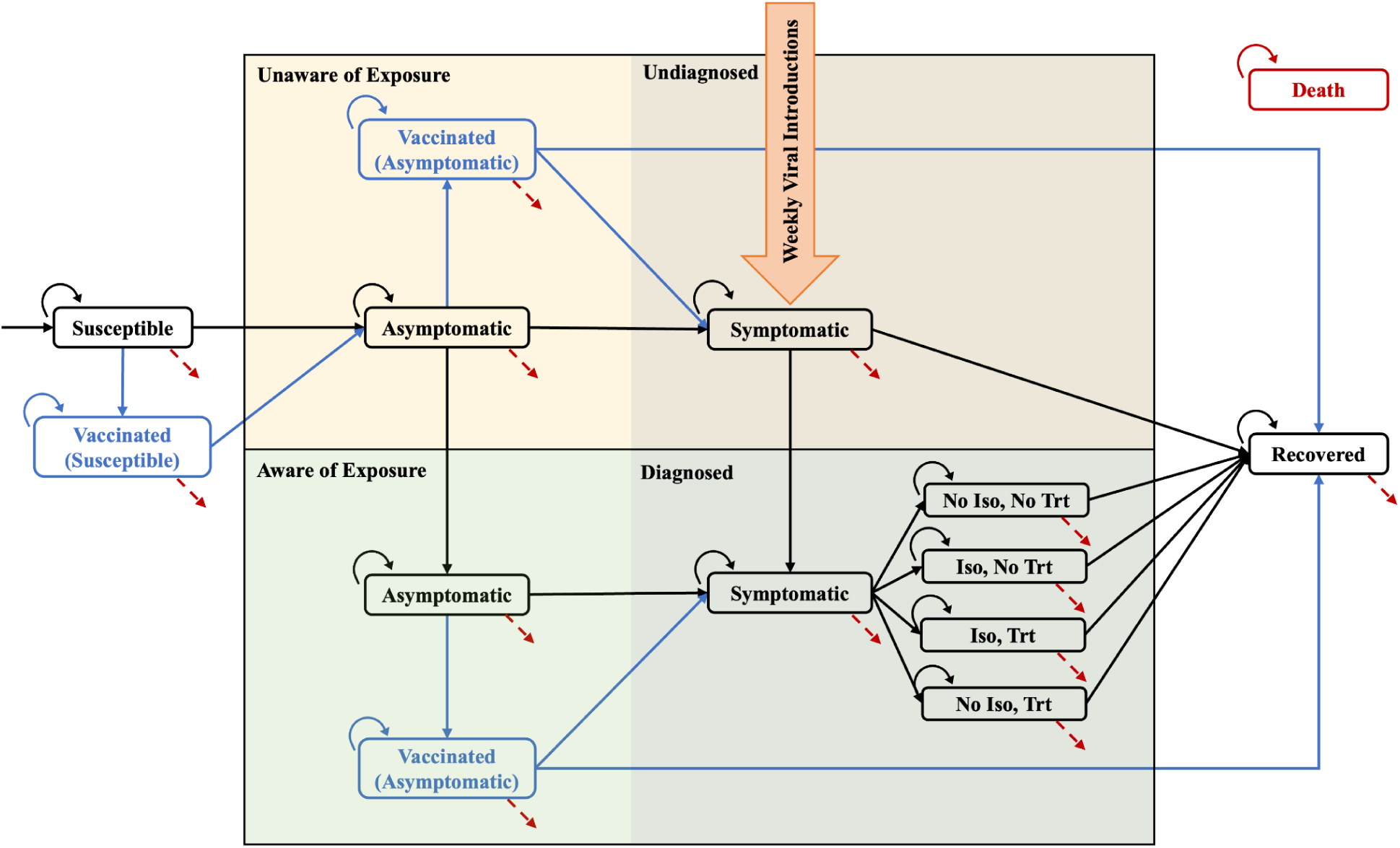
Schematic of the individual-based microsimulation model for mpox transmission dynamics in LAC with phylodynamics informed introductions. Our model simulates mpox transmission, vaccination, diagnosis, isolation, treatment, and recovery on a weekly cycle in LAC. Susceptible individuals may become infected and enter the asymptomatic phase before progressing to symptomatic illness, during which transmission occurs exclusively through sexual contact. Transmission risk is modulated by demographic-specific mixing patterns based on age and race/ethnicity, and individuals living with HIV are modeled as having higher susceptibility. Symptomatic individuals may be unaware or aware of exposure; awareness leads to earlier diagnosis and possible isolation and/or treatment. Preventive vaccination is available to susceptible individuals, while post-exposure prophylaxis can be administered to those recently exposed. Individuals transition to recovery or may exit the model through non-mpox-related death. New 15-year-old susceptible enter the model each week. Weekly viral introductions are incorporated as external symptomatic cases added directly into the undiagnosed symptomatic compartment. These cases contribute to transmission but are not counted as new diagnoses within LAC. Adapted from Liang et al (18)

**Figure S17:**
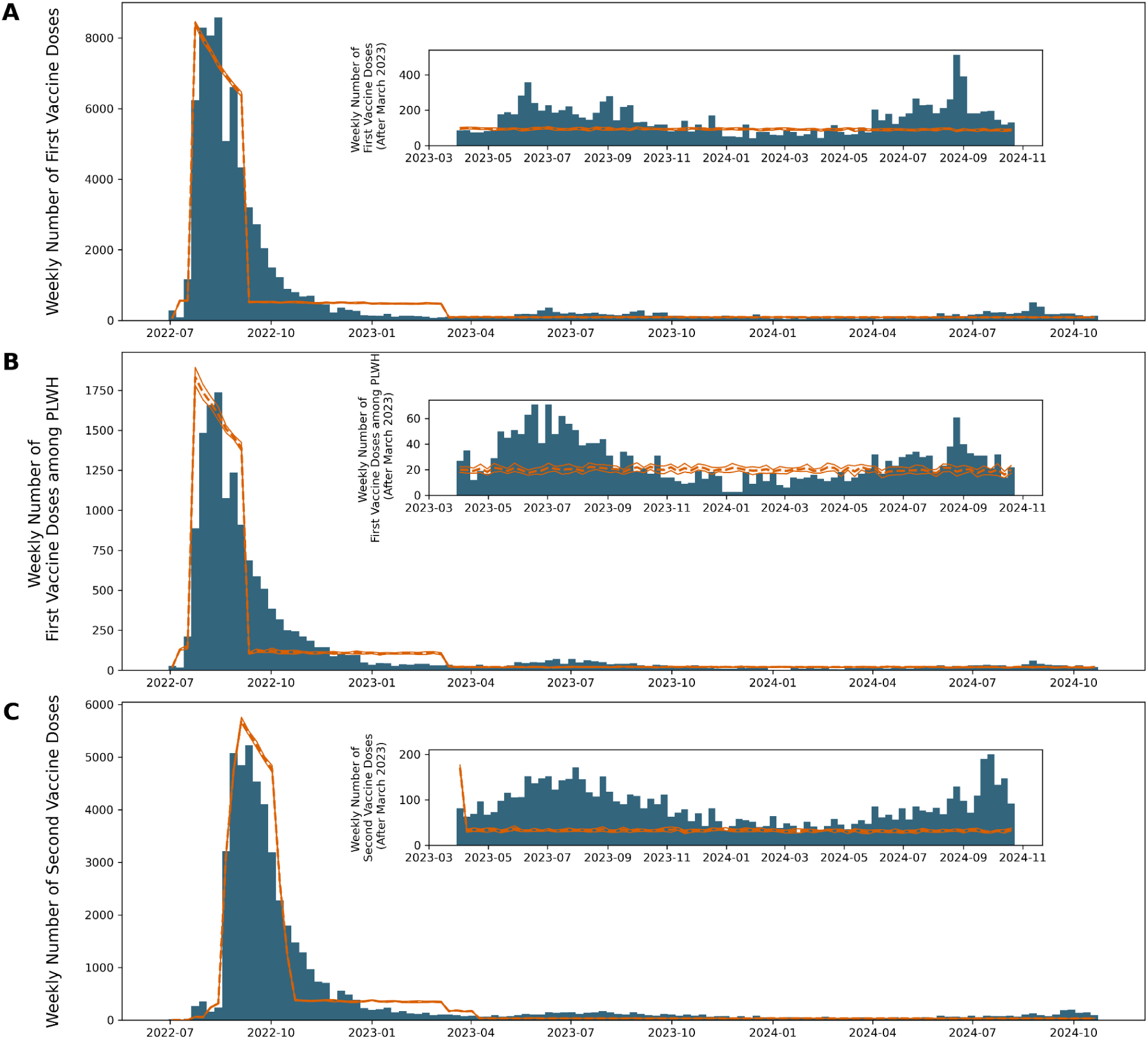
Vaccination validation of microsimulation model. All three panels represent the comparison between the empirical number of vaccination doses given (dark blue) and the number of doses administered as simulated by our model (orange). The dashed orange line represents the mean; the bands represent the 95% uncertainty interval calculated via bootstrapping. The inset graphs for each panel represents the same data but only after March 2023 to allow for better visualization of smaller numbers. Panel **A** represents the comparison of the number of first doses of the mpox vaccine given, panel **B** is for the number of first doses among people living with HIV (PLWH), and panel **C** is the number of second doses given.

